# Life course socio-economic position and healthy ageing: A systematic review of longitudinal studies

**DOI:** 10.1101/2025.10.22.25338545

**Authors:** Yisheng Ye, Chengxu Long, Kia-Chong Chua, Darío Moreno-Agostino, Matthew Prina

## Abstract

**Background:** Increasing health inequalities among older adults globally illustrate the urgent need for effective interventions. Socio-economic position (SEP) may affect healthy ageing through various life course mechanisms. However, longitudinal associations between life course SEP and healthy ageing as a multidimensional construct remain unclear.

**Methods:** We conducted a comprehensive systematic review of longitudinal studies investigating the associations between life course SEP indicators and multidimensional healthy ageing outcomes. A systematic literature search was conducted across four databases (MEDLINE, Embase, PsycINFO, and Web of Science) from inception to April 2025. Due to the heterogeneity in the operationalisation of SEP and healthy ageing, a narrative synthesis was performed (Prospero CRD42023418728).

**Results:** 47 articles were included in the review. Across multiple SEP indicators and life stages, higher educational levels (39/43 studies) and higher income/wealth (31/36 studies) were positively associated with better healthy ageing. Occupation showed inconsistent evidence. Life-course evidence showed childhood SEP disadvantage predicted poorer later-life outcomes (13/17 studies), with cumulative multi-stage disadvantage showing additive effects (5 studies) and upward mobility conferring benefits (3 studies). These patterns manifested in three age-related inequality trajectories: widening (18/23 studies), convergence, and persistence, with education-cognitive disparities showing strongest widening effects. Cross-national evidence revealed regional specificities. Studies also identified gender moderation effects (5 studies) and examined mediating pathways (6 studies).

**Conclusion:** Education showed the most consistent protective effects, while income/wealth effects were complex. Health inequalities widened with age, highlighting lasting childhood impacts. Targeted interventions addressing early educational investment and life stage-specific strategies are needed for reducing healthy ageing inequalities.

## 1. Introduction

The global population is ageing rapidly, with the population aged 60 and above projected to reach 2.1 billion by 2050 (United Nations, 2022). This demographic shift accentuates longstanding health inequalities, which are particularly pronounced among older adults (de la Fuente et al., 2019; Sadana et al., 2016). A key driver of these disparities is the accumulation of socio-economic differences experienced throughout an individual’s life course, leading to distinct health outcomes in later life (Cullati, 2015; Kuh et al., 2003; Liu et al., 2023a; Smith, 2007; Whitley et al., 2016). Research demonstrates that the gap in healthy life expectancy between different socio-economic status groups can span several years (Chen, 2024; Payne and Xu, 2022; Xu and and Payne, 2024). Without effective policy interventions, the continued exacerbation of these health inequalities will generate a series of adverse consequences: it will not only compromise the quality of life for older people but also pose significant challenges to the sustainable development of social care systems (Beard et al., 2016; Wu et al., 2020). Understanding how life course factors contribute to health inequalities has therefore become a critical research priority for promoting healthy ageing (Marengoni and Calderon-Larrañaga, 2020).

In response to these challenges, the concept of healthy ageing has evolved considerably. The conceptualisation has shifted from earlier models centred on static states, such as Rowe and Kahn’s emphasis on the ‘absence of major disease and disability’ (Rowe and Kahn, 1997), towards the World Health Organization’s (WHO) more dynamic framework, which defines healthy ageing as “an ongoing process of developing and maintaining the functional ability that enables well-being in older age” (WHO, 2015). Given the complexity of healthy ageing, disease-based concepts or those focused on single health domains may be insufficient to capture the full spectrum of health status (Behr et al., 2023; Tinetti and Fried, 2004). There is growing recognition that multidimensional approaches are needed, as different domains capture distinct but complementary aspects of overall health and well-being (Gao et al., 2022; Hsiao and Chen, 2024). This shift requires us to focus not only on health status itself, but also to understand the factors and mechanisms that shape health development over the life course.

A life-course approach provides a theoretical framework for exploring the causes of these dynamic processes. Different models within this framework can be helpful to examine the role of socio-economic position (SEP) on health and healthy ageing across and at different life stages (Mishra et al., 2010). For instance, the long-term effects of early-life factors such as SEP during specific sensitive periods on health trajectories in later life (the sensitive period model), the cumulative impact of SEP across different life stages (the accumulation model), and the influences of social mobility (the social mobility model) (Kuh et al., 2003; Wagner et al., 2024). These different theoretical models offer multiple perspectives for understanding the dynamic nature of health inequalities and highlight the need for longitudinal study designs to track individuals’ long-term health trajectories.

To date, only one systematic review has examined the relationship between SEP and healthy ageing (Wagg et al., 2021). This review combined cross-sectional and longitudinal studies but focused primarily on documenting overall associations rather than examining life course mechanisms. Moreover, their findings were based on evidence available up to February 2021, with most studies originating from high-income countries, which limits the global applicability of the findings. Research literature since then has expanded and deepened in several aspects. Specifically, three key developments can be identified: first, research focus has increasingly shifted from assessing associations at specific single time points to using longitudinal data to explore how SEP influences entire health trajectories (Chang et al., 2023; Guo et al., 2025; Wang et al., 2024). Second, analytical methods have become more sophisticated, with a growing number of studies investigating complex mechanisms such as social mobility, cumulative effects, and mediation analysis (Huang et al., 2025; Payne and Xu, 2022; Yu-Tzu et al., 2025). Finally, the evidence base has been broadened, with research findings from large cohorts in low- and middle-income countries (LMICs) such as China and Mexico, providing a broader global perspective for understanding patterns of health inequality across different social contexts (Salinas-Rodríguez et al., 2024; Zhou et al., 2024). Despite this growing interest, no systematic review has comprehensively examined the longitudinal associations between SEP and healthy ageing across the life course.

To address this gap and inform policy interventions for healthy ageing, we conducted a systematic review addressing the following research questions: (1) What is the strength and consistency of associations between different SEP indicators (e.g., education, income/wealth, occupation) and healthy ageing outcomes? (2) From a life course perspective, how does SEP across different life stages (e.g., childhood, adulthood) and through different patterns (e.g., accumulation, social mobility) influence the dynamic processes of healthy ageing? (3) Do the patterns and mechanisms of these associations vary according to macro social contexts (e.g., national income levels) and individual characteristics (e.g., gender)?

## 2. Methods

The protocol for this systematic review was prospectively registered with the International Prospective Register of Systematic Reviews (PROSPERO) (registration number: CRD42023418728). Study execution followed the pre-registered protocol, with an updated search round to capture recent literature.

The report follows the guidance of the Preferred Reporting Items for Systematic Reviews and Meta-Analyses (PRISMA) 2020 statement (Page et al., 2021), with additional reference to the PRISMA-Equity extension (Welch et al., 2015). A PRISMA checklist is attached in supplementary material 1.

### 2.1 Eligibility Criteria and Search Strategy

Detailed inclusion and exclusion criteria for this review are presented in supplementary material 2. In brief, we included longitudinal studies that: (a) recruited community-dwelling, middle-aged or older adults (≥50 years) at baseline; (b) assessed SEP during at least one life-course stage; and (c) measured healthy ageing using a composite index integrating at least two health domains in order to more appropriately capture the multidimensionality of the healthy ageing concept, as single-domain measures (for example, physical or cognitive function alone) may inadequately reflect this complexity and fail to provide a comprehensive understanding of healthy ageing in older adults.

A systematic literature search was conducted on Medline (via PubMed), Embase (via Ovid), PsycINFO (via Ovid), and Web of Science Core Collection. The search strategy combined free-text terms and subject headings (e.g., MeSH), structured around three core concepts: (1) SEP; (2) healthy ageing; and (3) life course and longitudinal design. The complete search strategy is provided in supplementary material 2. Initial searches covered database inception to 25 March 2023, with updated searches conducted in April 2025. Searches had no language restrictions, but only English-language studies were included. Reference lists of all included studies were also hand-searched to identify any relevant articles missed by electronic searches.

### 2.2 Study Screening

Study screening was conducted in two stages independently by two reviewers. A calibration exercise was first performed on 10 randomly selected articles, achieving 60% initial agreement and 100% agreement after discussion. Using the Rayyan platform, reviewers independently screened titles and abstracts of all retrieved articles. Potentially eligible studies proceeded to full-text assessment, again conducted independently by two reviewers. At both stages, articles deemed potentially relevant by either reviewer were retained. Disagreements were resolved through discussion or by a third researcher who acted as an arbiter if consensus could not be reached. Reasons for exclusion were documented at the full-text stage.

### 2.3 Data Extraction

Two researchers independently extracted data using a standardised data extraction form (Microsoft Excel). Any discrepancies were resolved through discussion, with arbitration by a third researcher when necessary.

Information was extracted on the following aspects:

1. Study characteristics: citation details, country of study and its World Bank income level classification, cohort name, and study design and follow-up period.
2. Population characteristics: baseline characteristics such as sample size, age, sex composition, and response rate.
3. SEP measurement: type of indicator, timing of measurement (e.g., childhood, adulthood), and operationalisation.
4. Healthy ageing measurement: the conceptual framework used (e.g., intrinsic capacity, frailty index), the health domains included, and the specific measurement method (e.g., dichotomous, continuous score, or categorical profiles derived from latent class analysis).
5. Analytical strategy: the statistical models employed, key covariates controlled for in the models, methods for handling missing data, and whether stratified analyses were performed.
6. Main results: the direction of the association between SEP and healthy ageing, adjusted effect estimates (e.g., odds ratios, beta coefficients, and their 95% confidence intervals), and the authors’ main conclusions.

### 2.4 Quality Assessment

We used the Joanna Briggs Institute (JBI) Critical Appraisal Checklist for Cohort Studies to assess the methodological quality of each included study (Barker et al., 2025). This tool comprises 11 items designed to systematically evaluate the risk of bias in areas such as participant selection, exposure and outcome measurement, control of confounding factors, and follow-up.

Two researchers independently assessed each study, judging each item as ‘Yes’, ‘No’, ‘Unclear’, or ‘Not Applicable’. Discrepancies in the assessment were resolved through discussion or by consulting a third, senior researcher.

We also noted potential issues with ‘Table 2 Fallacy’, where adjustment variable coefficients from models designed to estimate the main exposure effect are inappropriately interpreted as independent causal effects (Westreich and Greenland, 2013).The assessment of Table 2 Fallacy was particularly relevant as SEP is often adjusted for rather than studied as the main exposure, increasing this risk. The results of the quality assessment were used in the sensitivity analysis and interpretation of findings; no studies were excluded based on their quality assessment.

### 2.5 Data Synthesis

Through systematic evidence mapping of the included studies, we identified significant methodological heterogeneity in the operationalisation of healthy ageing, the timing and type of SEP indicators measured, and the statistical methods employed (Althuis et al., 2014). A quantitative meta-analysis was therefore considered inappropriate.

Narrative synthesis was used to synthesise data (Popay et al., 2006). As no meta-analysis was conducted, items relating to quantitative synthesis (such as sensitivity analyses, heterogeneity exploration, and certainty assessment) were not applicable. In line with the review’s core questions, we systematically summarised and presented the findings organised by the different SEP indicators and conceptual frameworks for healthy ageing. During the synthesis, we paid particular attention to consistent patterns and variations in the findings across studies.

## 3. Results

The initial database search identified 7,455 records. After the removal of 3,200 duplicates, the titles and abstracts of the remaining 4,255 records were screened. At this stage, 4,193 records were excluded for not being relevant to the research topic, leaving 62 articles for full-text assessment. Following a detailed review of these 62 full-text articles, 30 were excluded for failing to meet the inclusion criteria. The most common reasons for exclusion were the reporting of a single dimension of healthy ageing (n = 12) or an ineligible study design (e.g., cross-sectional study, n = 8). The initial search therefore yielded 32 studies.

In the updated search, conducted in April 2025, an additional 1,757 records were identified. After removing 549 duplicates, 1,208 records underwent title and abstract screening, of which 1,173 were excluded. This left 35 articles for full-text screening, from which 20 were excluded. Similar to the initial search, the most frequent reasons for exclusion were the reporting of a single dimension of healthy ageing (n = 7), an ineligible study design (n = 5), or irrelevance to the research questions (n = 5). This updated search yielded 15 additional studies.

Combining both search rounds, a total of 47 studies were included in this systematic review. The complete study selection process is illustrated in the PRISMA flow diagram in Figure 1.

**Figure 1:**
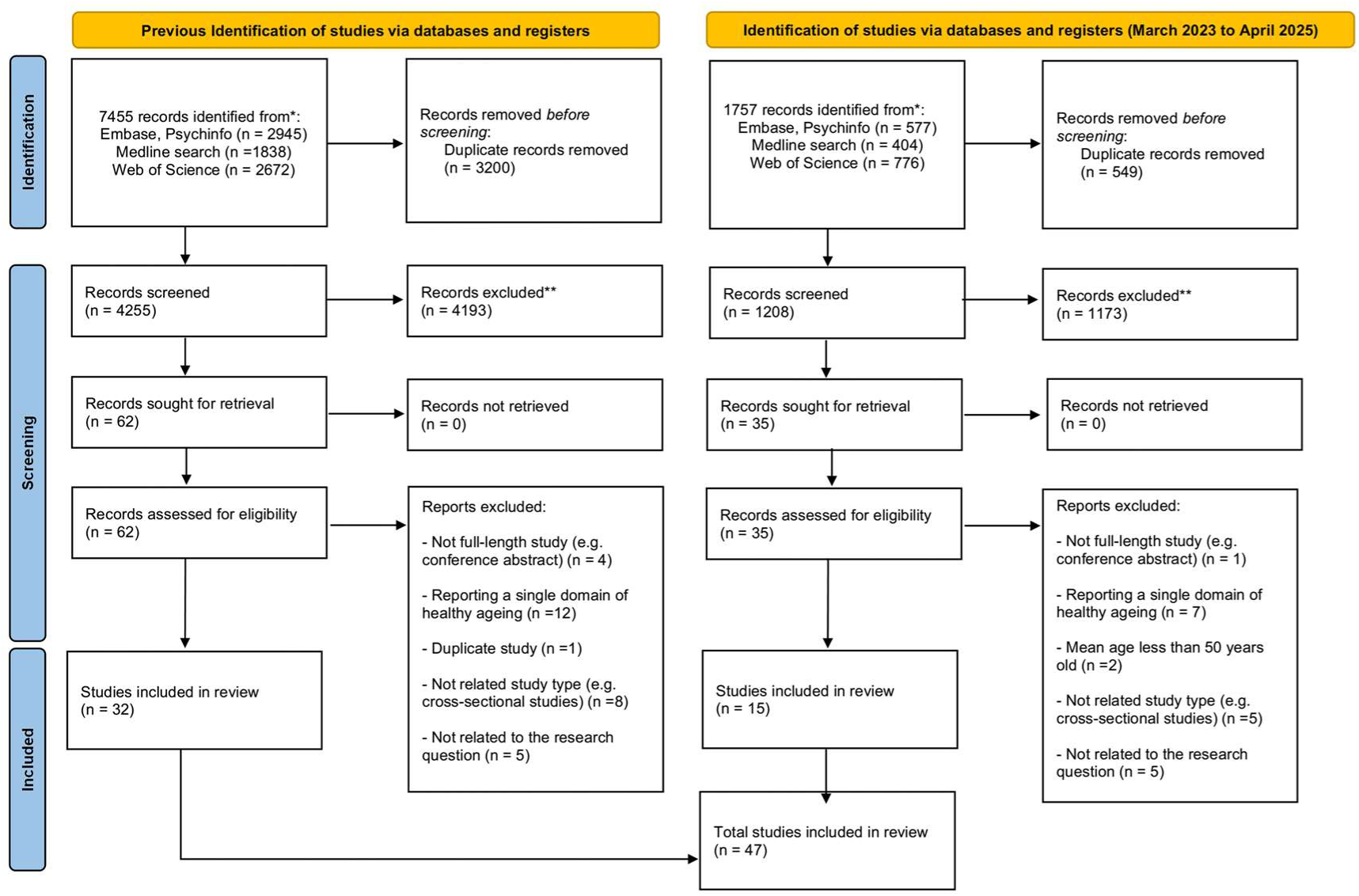
PRISMA flowchart

### 3.1 Characteristics of Included Studies

The detailed characteristics of the 47 included longitudinal studies are summarised in Table 1. These studies exhibited considerable diversity in terms of geographical distribution, time span, population characteristics, and methodological quality.

**Table 1.**
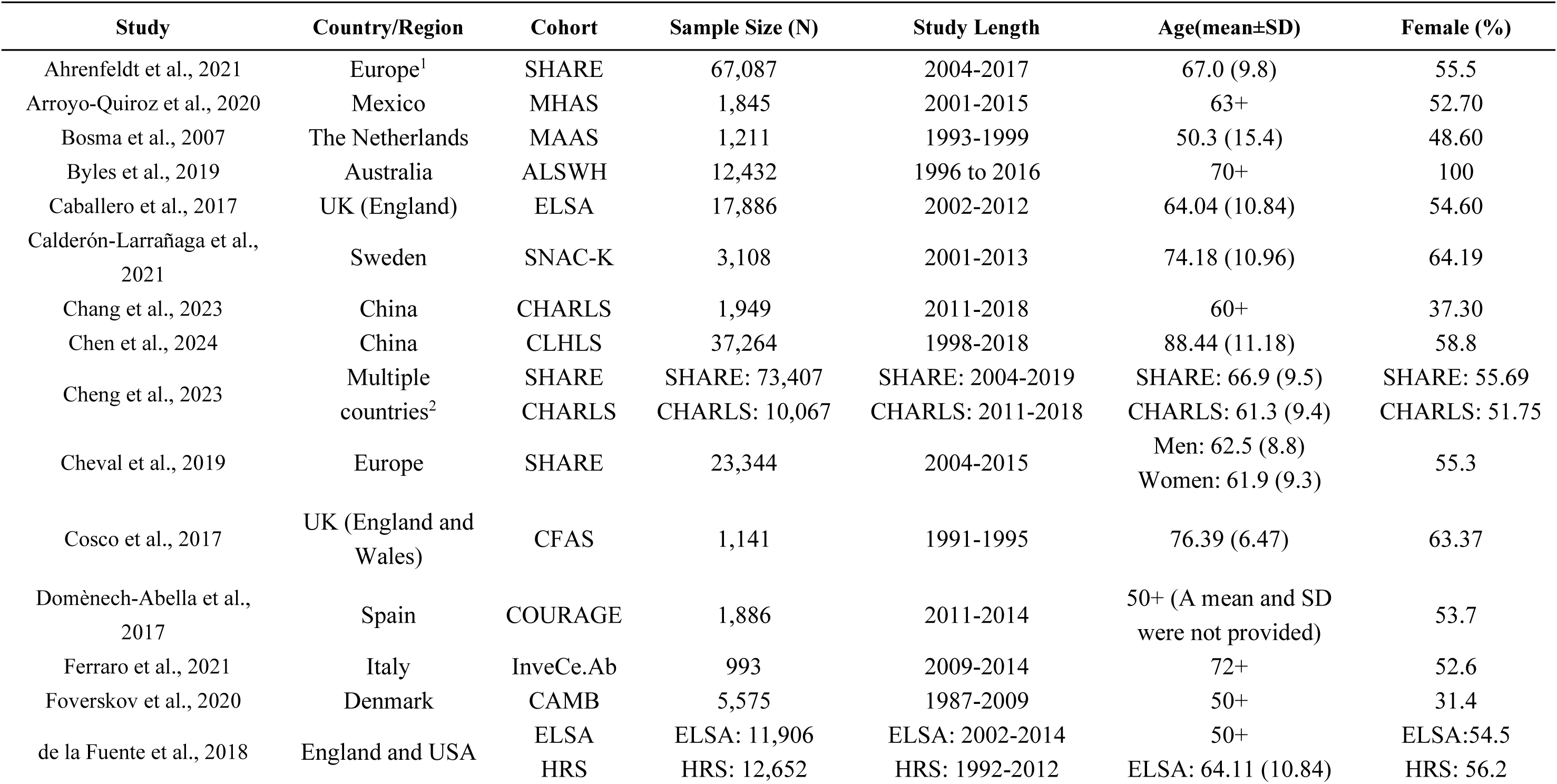
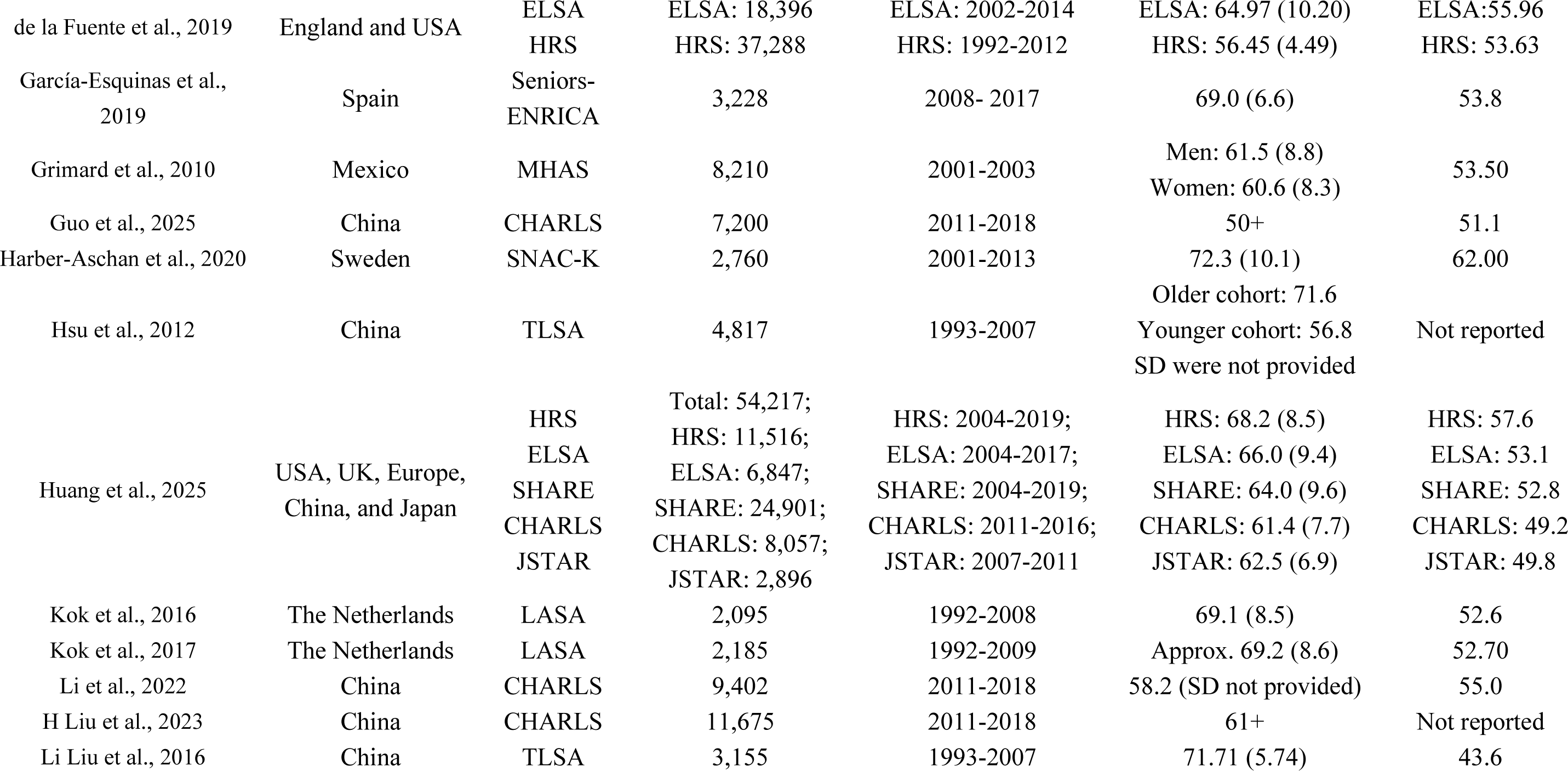
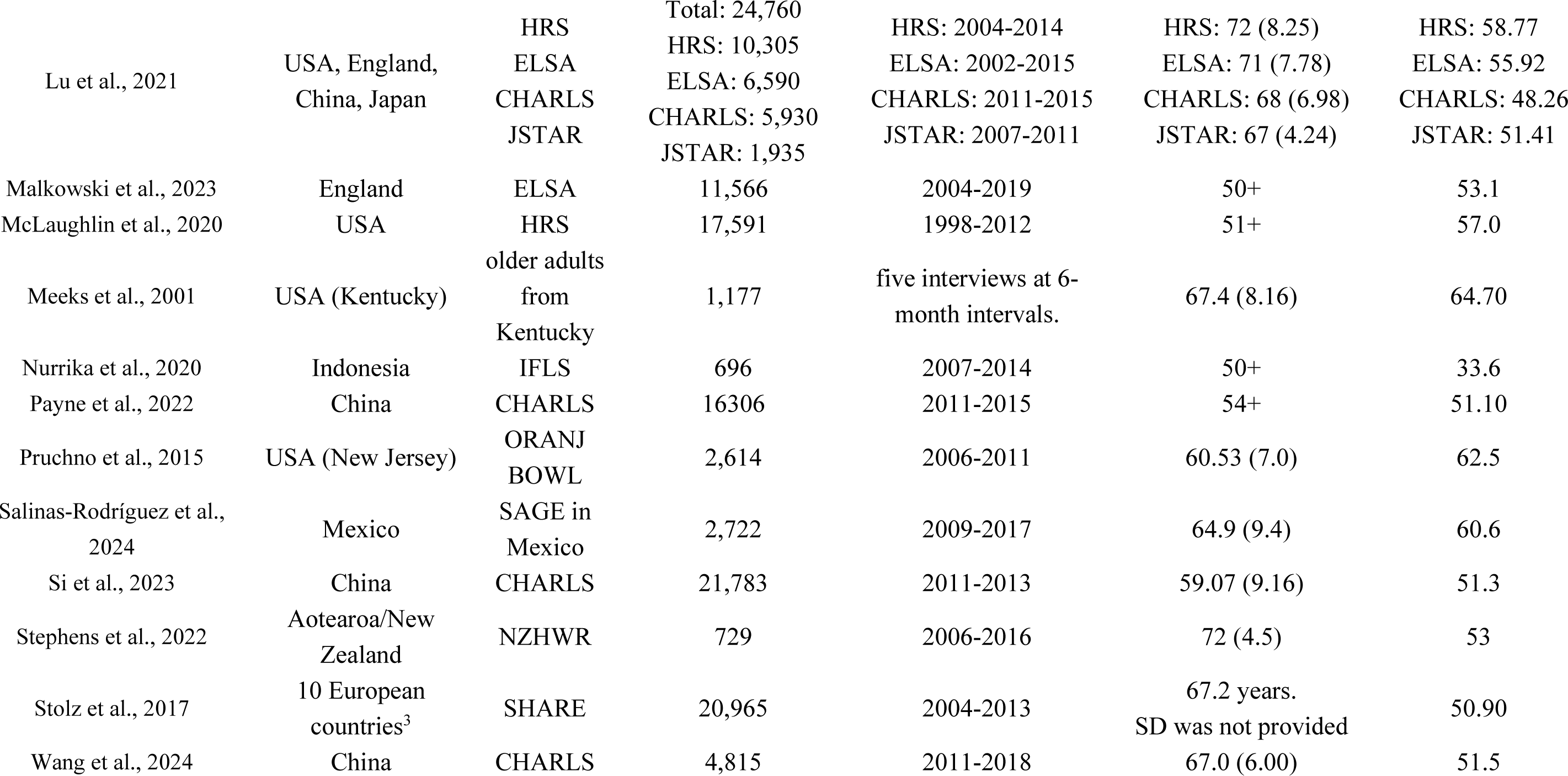
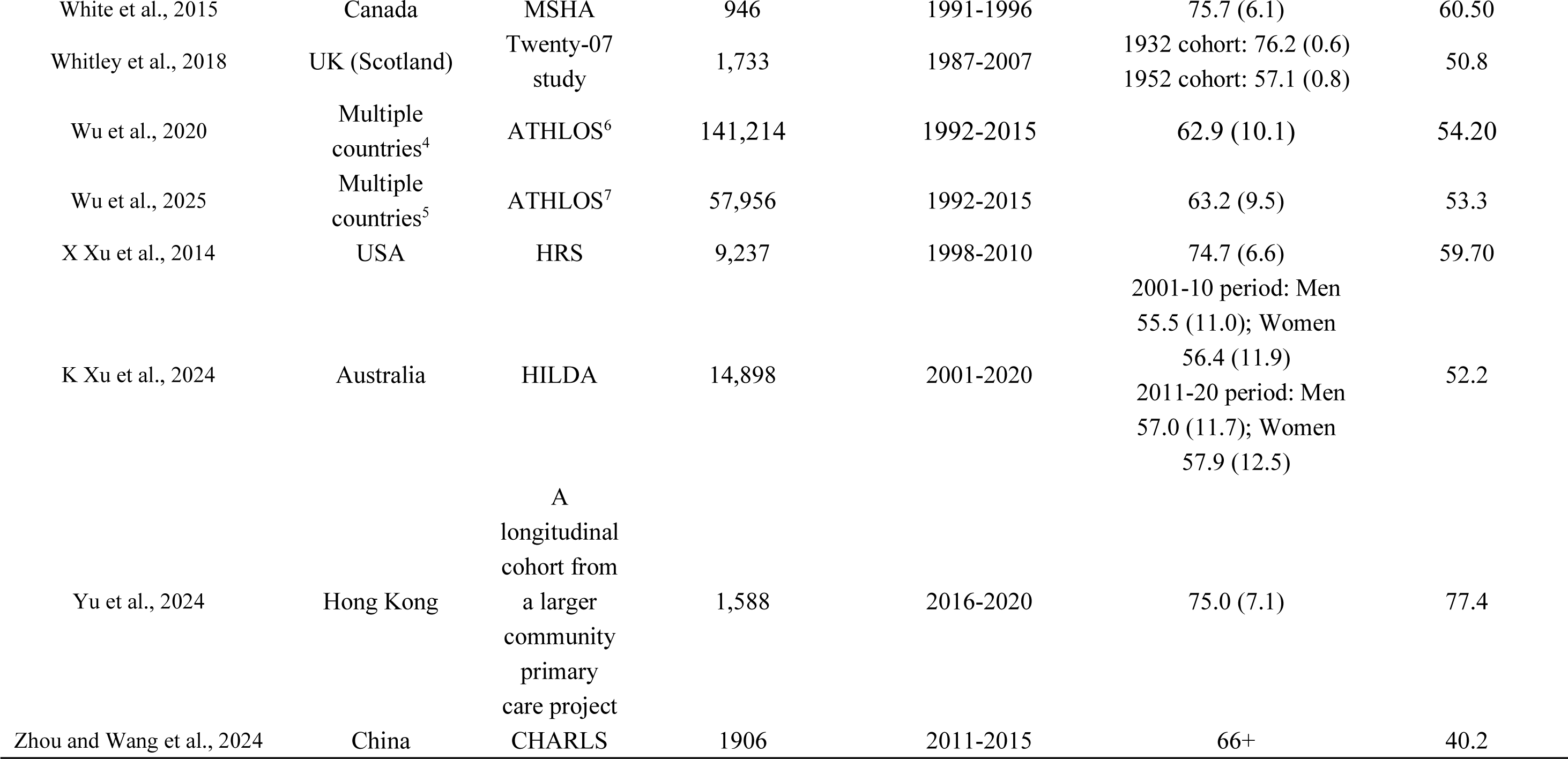
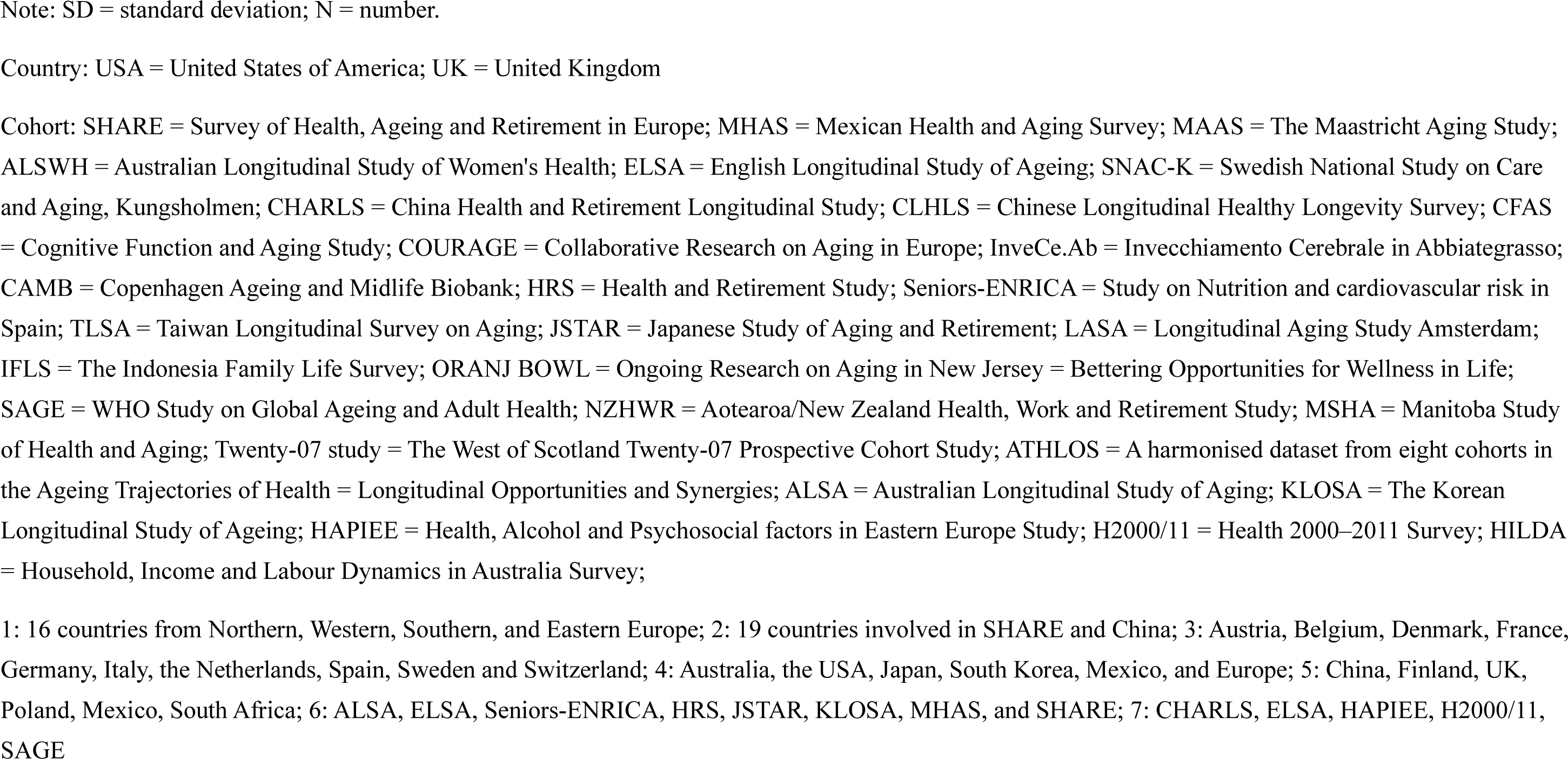
Characteristics of included longitudinal studies.

Risk of bias assessment with the JBI Critical Appraisal Checklists revealed the following sources of bias: unclear measurement of exposure validity and reliability in seven (15%) studies; no reporting of whether participants were free of the outcome at baseline in five (11%) studies; unclear measurement of outcomes validity and reliability in two (4%) studies; unclear reporting of follow-up time adequacy in one (2%) study; incomplete follow-up or inadequate description of loss to follow-up reasons in fifteen (32%) studies; and the absence of strategies to address incomplete follow-up in eleven (23%) studies.

Figure 2 shows the risk of bias summary plot. The details of each study are provided in the supplementary material 3.

**Figure 2:**
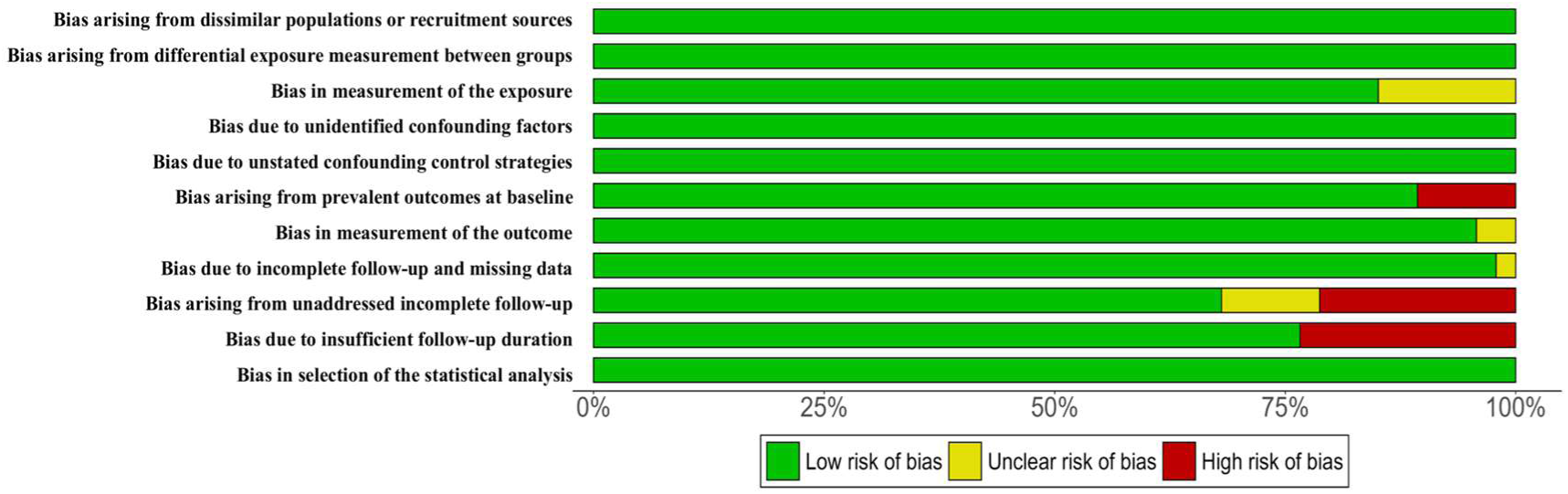
Risk of bias summary plot

### 3.1.1 Geographical Distribution

Of the 47 studies, 25 (53.2%) were from high-income regions such as North America, Europe, and Australia, while 22 (46.8%) included samples from low- and middle-income countries (LMICs). Among these 22 studies, 15 were based entirely on LMIC populations, and 7 were cross-national comparative studies. The LMIC studies were predominantly from mainland China (n = 11), Mexico (n = 3), and Indonesia (n = 1).

#### 3.1.2 Publication Date and Follow-up Duration

The publication years of the included studies ranged from 2001 to 2025. Of these, 17 studies (36.2%) were published in 2022 or later, and 30 of 47 studies (63.8%) were not included in the previous systematic review. Follow-up duration ranged from 2 to over 20 years.

#### 3.1.3 Population Characteristics

All studies recruited community-dwelling adults aged 50 years or older at baseline. The reported mean age at baseline ranged from 50.3 to 88.4 years, with most studies focusing on a baseline age range of 55–65 years. Regarding sex composition, all but one study, which focused exclusively on women (Byles et al., 2019), included both male and female participants, with the proportion of women ranging from 31.4% to 77.4%.

Study size varied significantly, with sample sizes ranging from 696 (Nurrika et al., 2020) to over 140,000 participants (Wu et al., 2020). 26 studies (55.3%) had a sample size of more than 5,000.

### 3.2 Overview of Measurement Methods for Healthy Ageing and SEP

The 47 included longitudinal studies showed significant methodological heterogeneity in their measurement of healthy ageing and SEP. As some studies employed multiple measurement methods or theoretical frameworks, the frequencies for the following categories may overlap. The details can be seen in supplementary material 4.

The detailed findings are presented in Table 2 and Table 3, organized by the life course coverage of SEP measures: Table 2 shows studies with SEP measures covering a single life stage in relation to healthy ageing measures across multiple time-points, while Table 3 shows studies with SEP measures covering multiple life stages in relation to healthy ageing measures across single or multiple time-points.

**Table 2.**
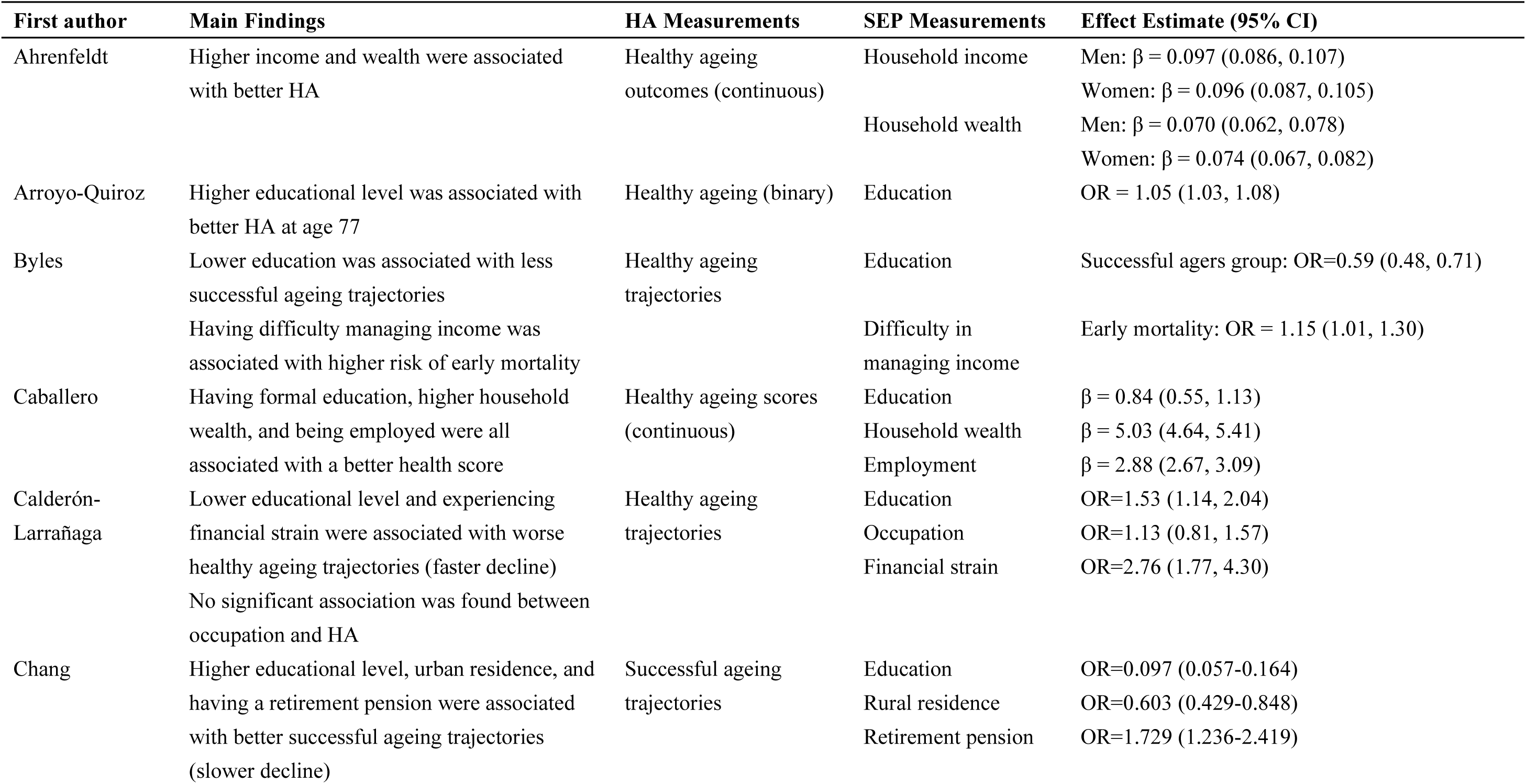
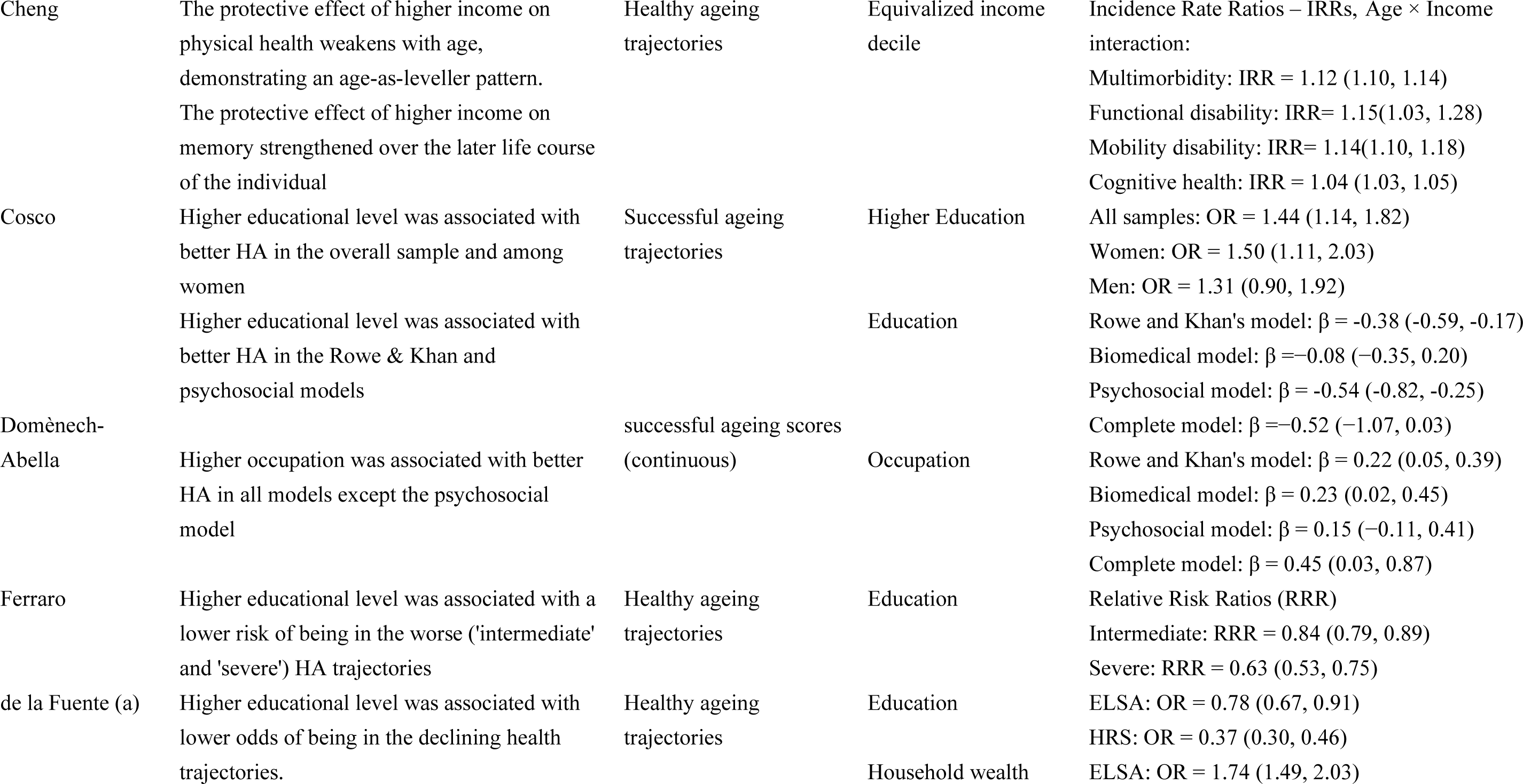
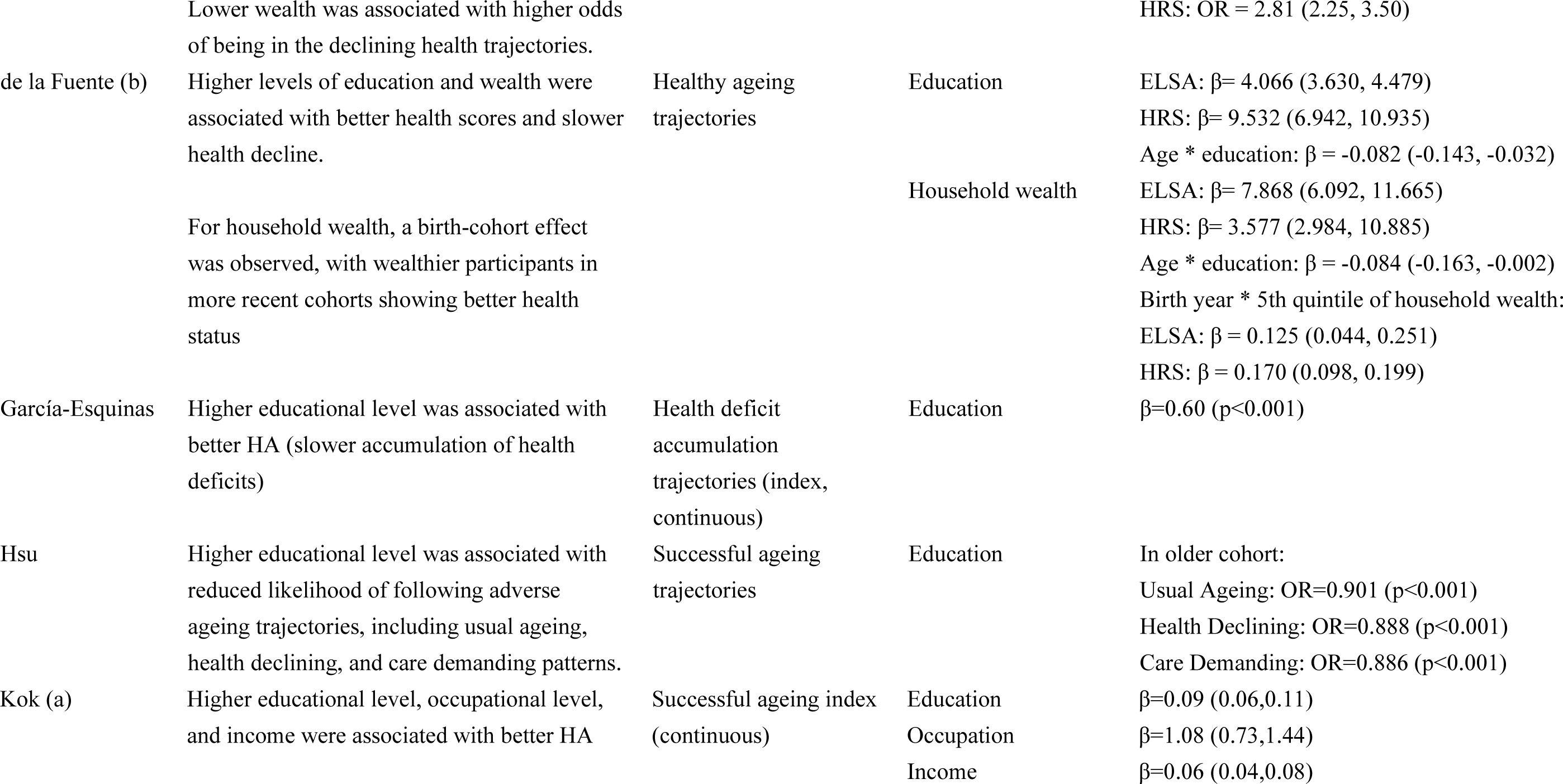
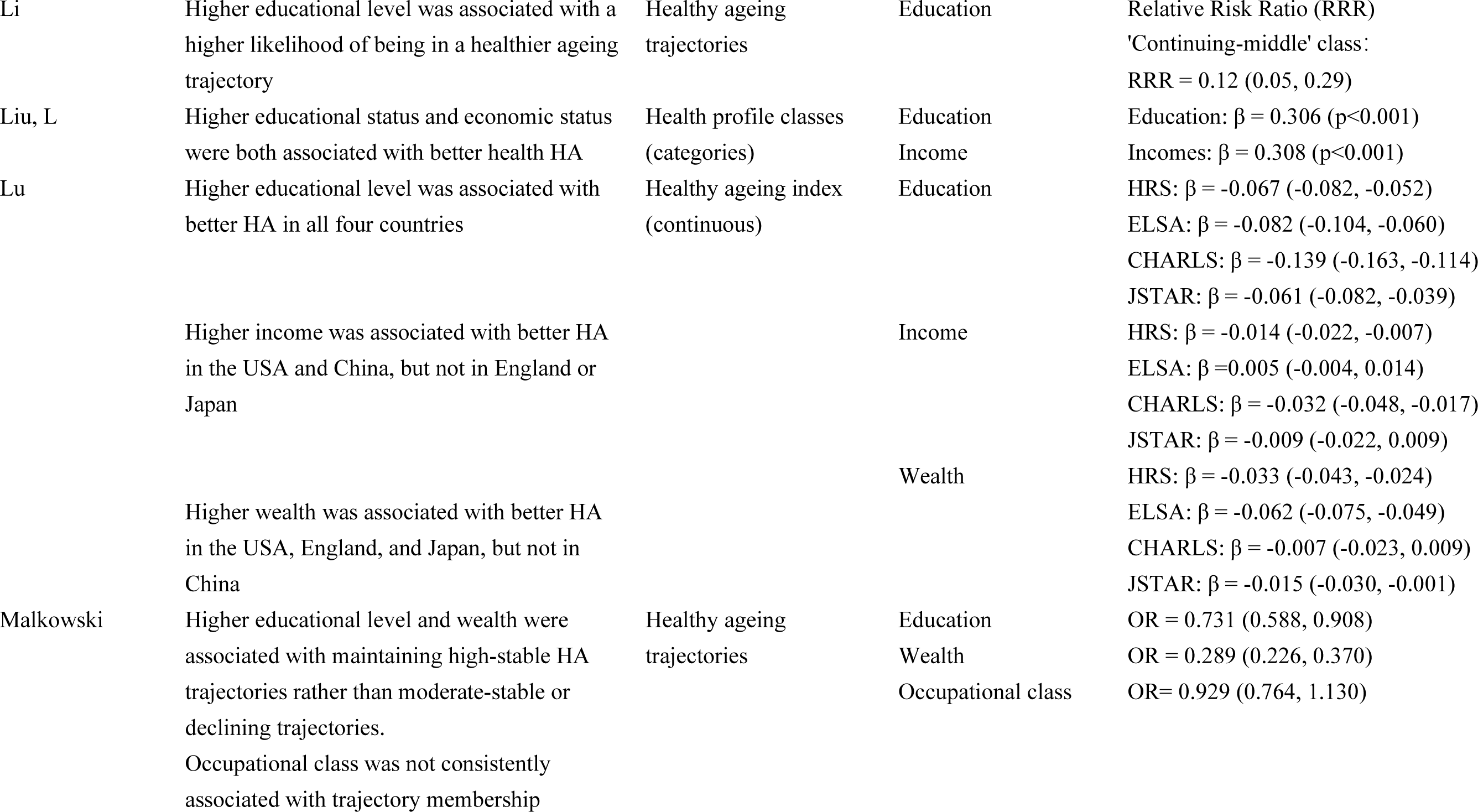
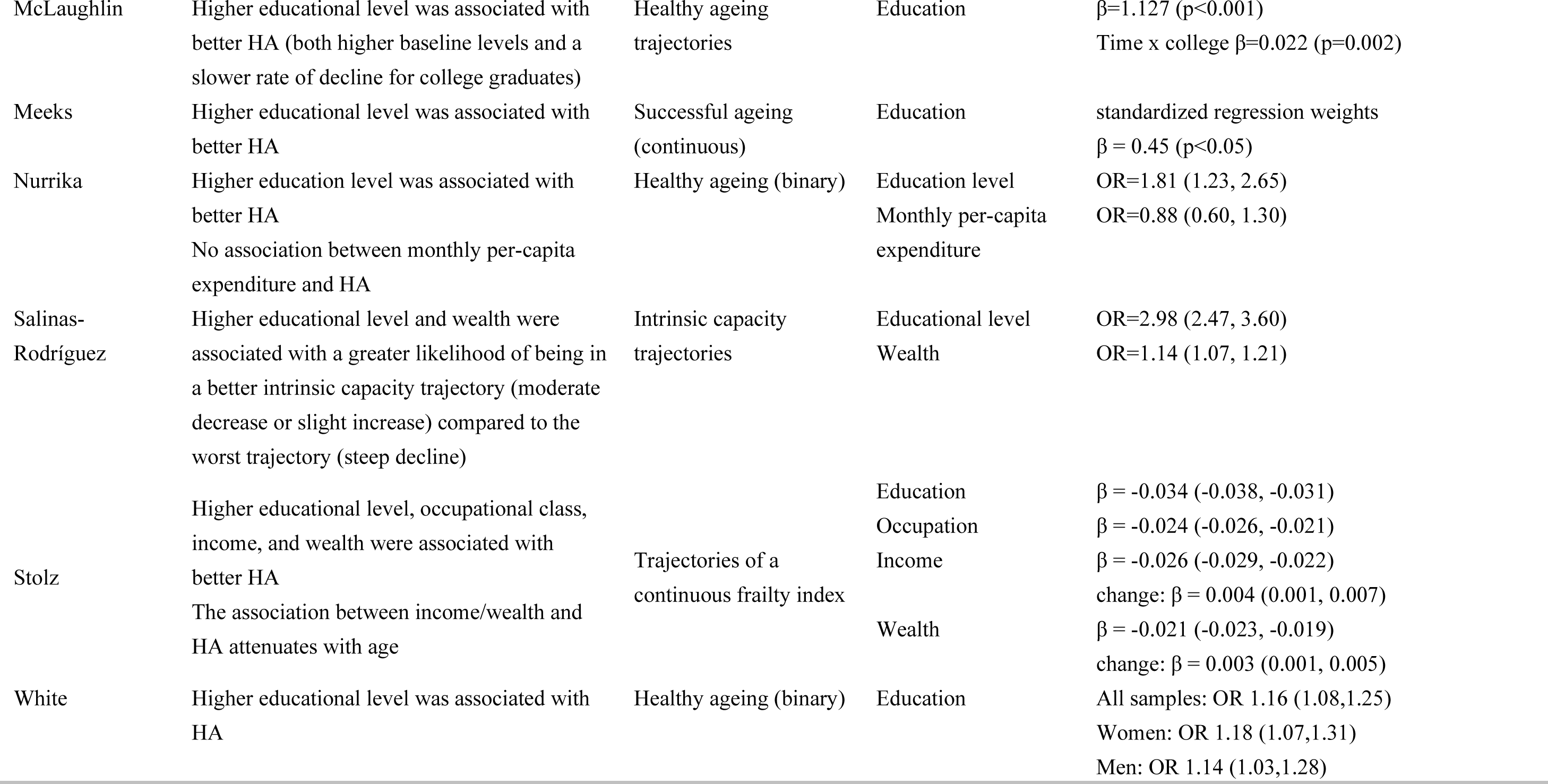
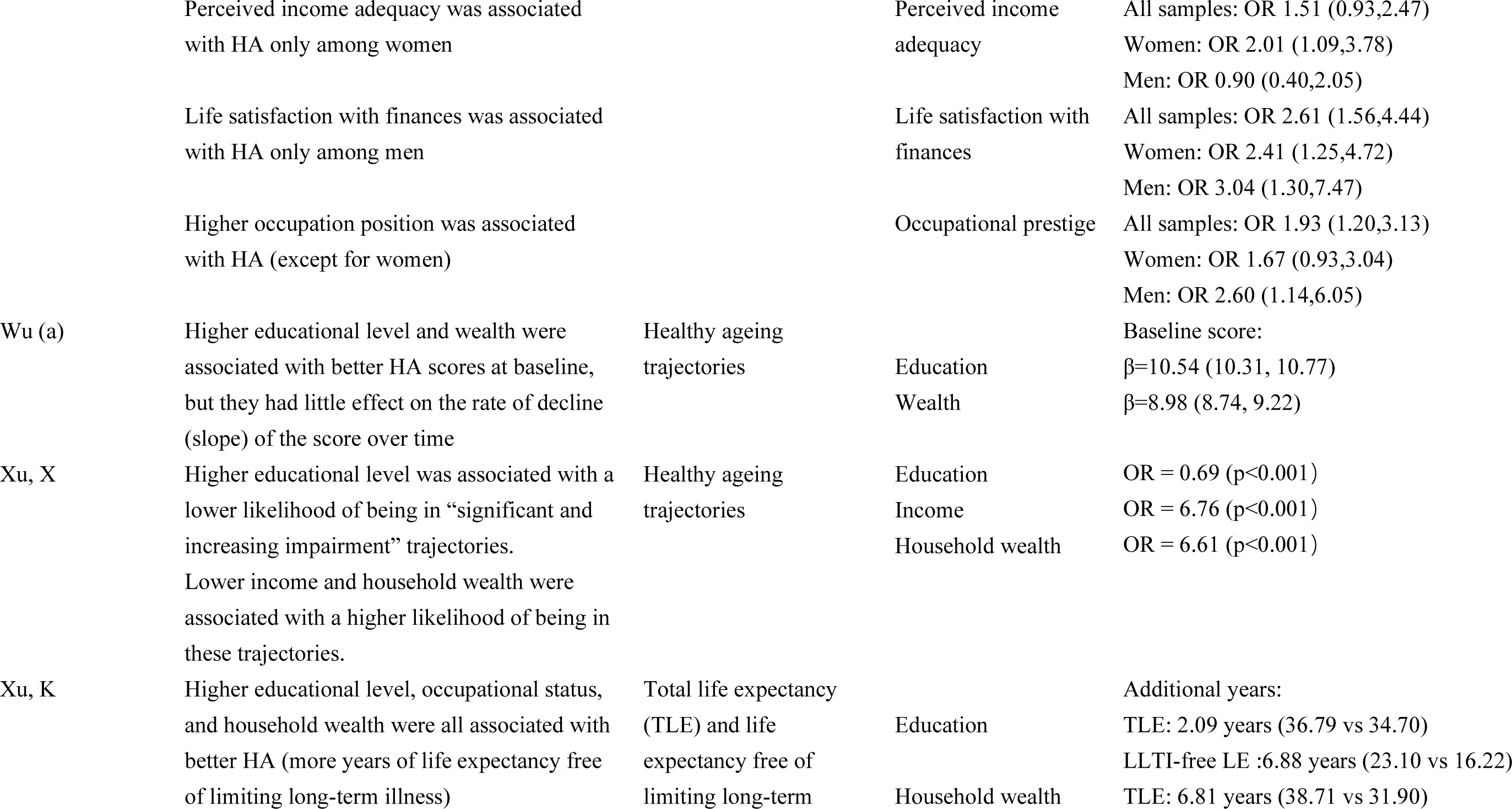
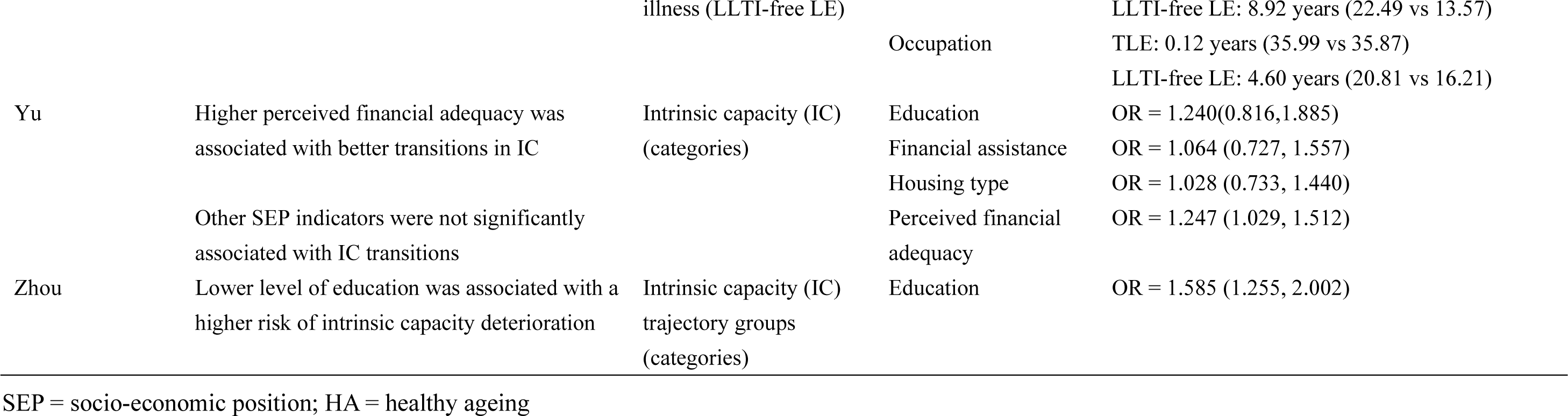
Association between SEP and healthy ageing outcomes: Single life-stage SEP measures.

**Table 3.**
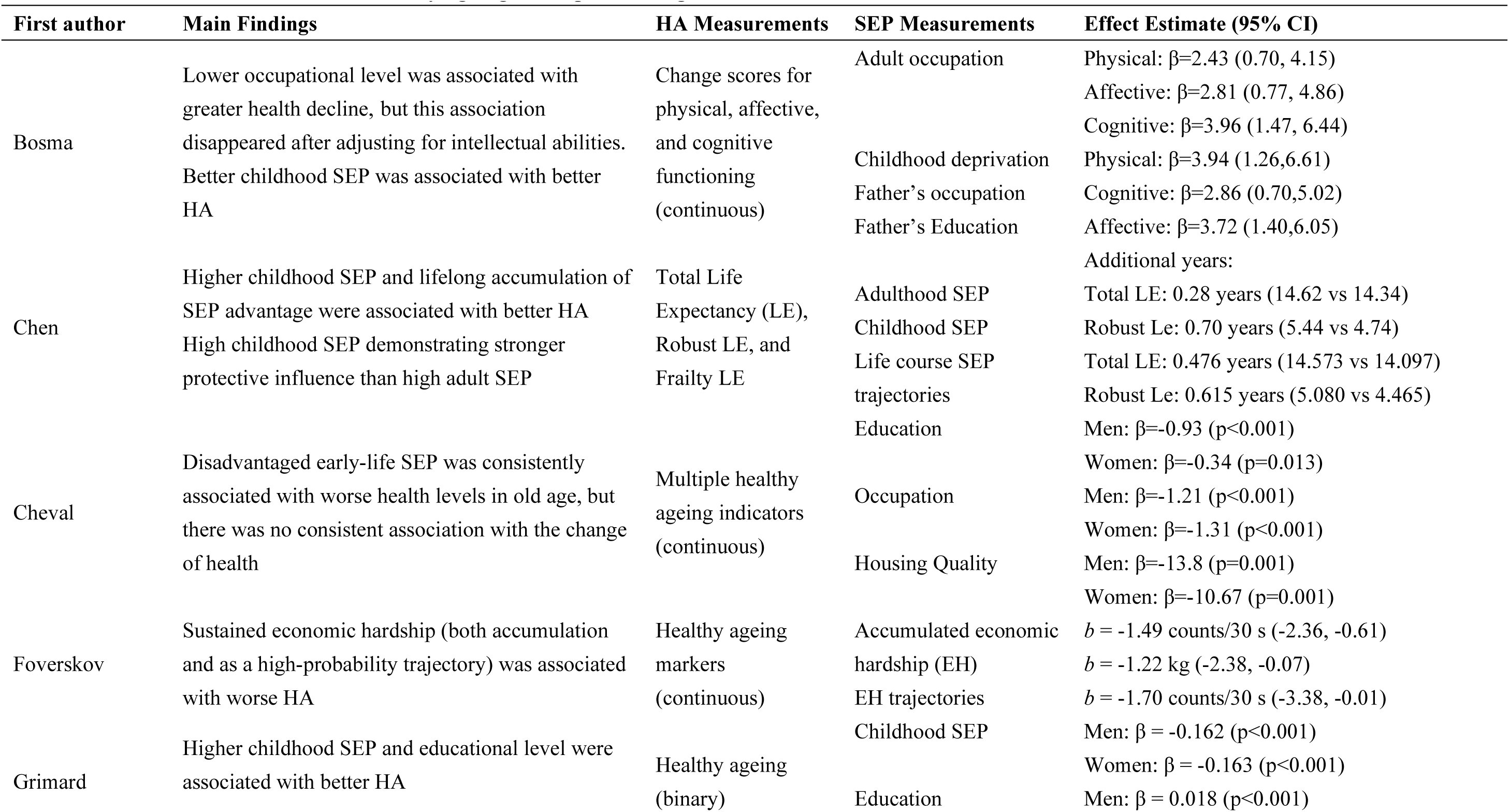
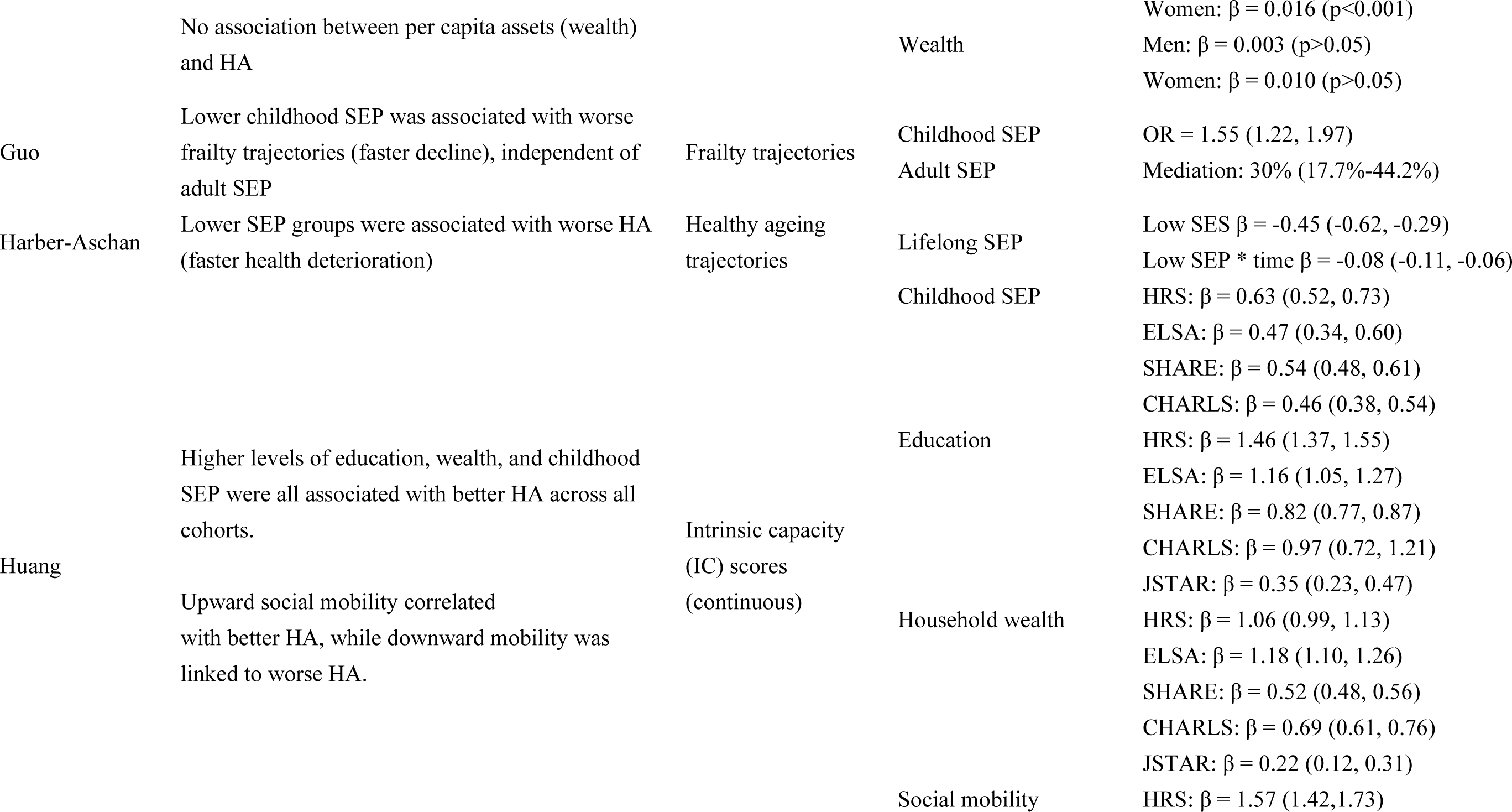
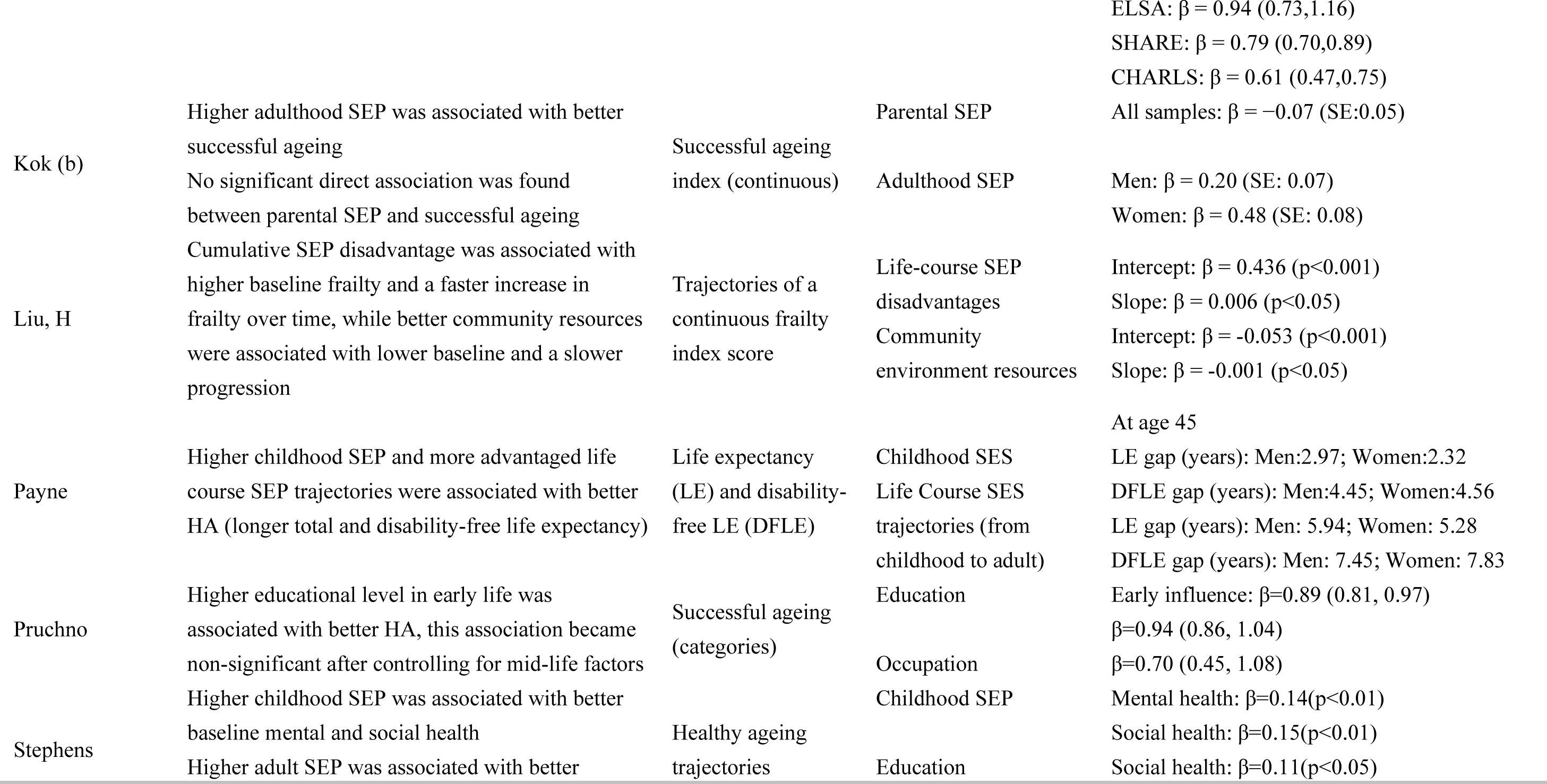
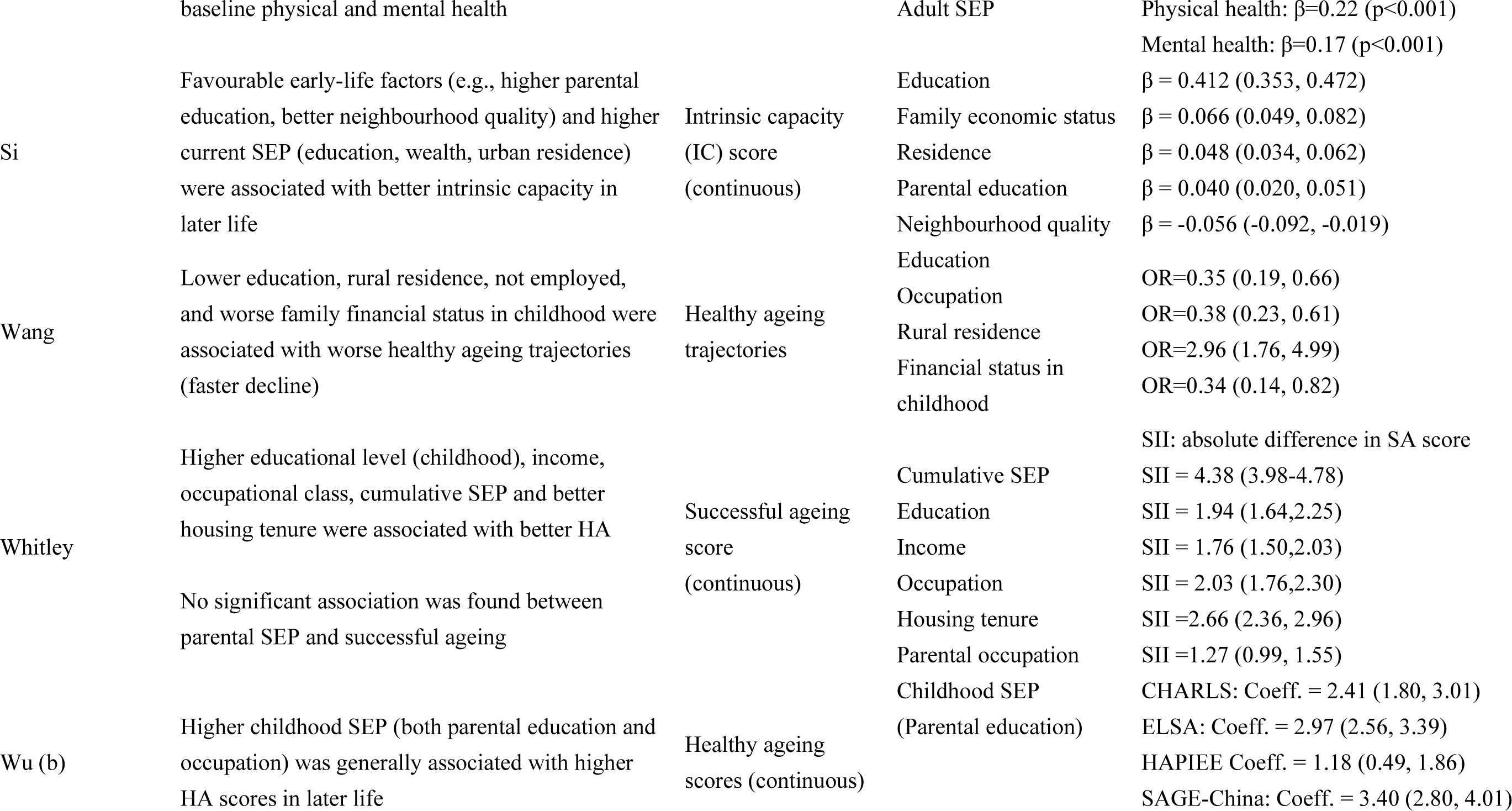
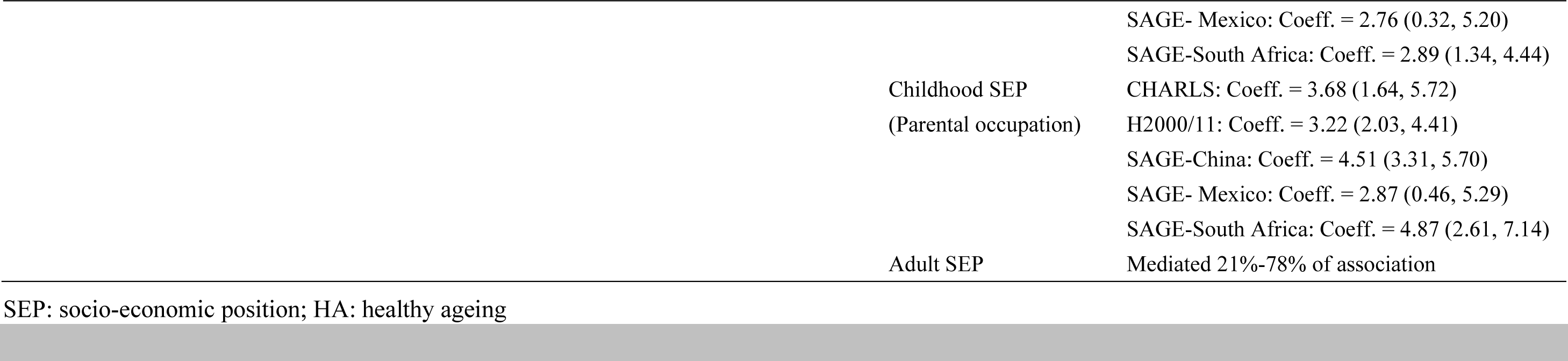
Association between SEP and healthy ageing: Multiple life-stage SEP measures.

#### 3.2.1 Measurement of Healthy Ageing

Researchers used a variety of theoretical models to conceptualise healthy ageing. The most common approach was developing new study-specific models (n = 26). Of the established theoretical frameworks, the WHO intrinsic capacity model (n = 7), the Rowe & Kahn successful ageing model (n = 5), and the models based on a frailty index or health deficit accumulation (n = 4) were the three most applied. In addition, 3 studies employed biomedical model, 2 studies used a health expectancy approach.

Multiple health dimensions were incorporated across studies to measure healthy ageing. The most represented health dimensions were physical function (n = 47, all studies), followed by cognitive function (n = 37), mental health (n = 25), personal perception (n = 23), and absence of disease (n = 19).

As physical function appeared in all studies, combinations including it merely reflected the frequencies of the other dimensions individually. Therefore, the analysis focused on the remaining dimensions. The highest combination frequency was found for cognitive function and mental health (n = 23), followed by cognitive function with personal perception (n = 18) and mental health with personal perception (n = 16). Notably, 92% of studies that included mental health also incorporated cognitive functioning, while 14 studies examined cognitive functioning independently. The most common three-dimensional combination was cognitive function, mental health, and personal perception (n = 15). Combinations involving healthy survival were rare, appearing in only 3 studies.

Regarding the operationalisation of the healthy ageing outcome, studies used different approaches: trajectory analysis to identify distinct health patterns over time (n = 21), continuous scores (n = 18), binary variables (n = 4), and multi-categorical variables (n = 4).

#### 3.2.2 Measurement of SEP

The SEP measurements in the included studies reflected different life course perspectives. Most studies (n = 30) measured SEP at a single life stage, while 17 studies measured SEP across multiple life stages, with early-life indicators typically obtained through retrospective reporting. Among these 17 studies, seven constructed specific life course SEP indicators. (e.g., trajectory patterns, cumulative scoring, social mobility indicators).

For specific SEP indicators, education (n = 43), income or wealth (n = 36), and occupation (n = 22) were the three most used dimensions, with most studies (n = 35) examining at least two indicators concurrently.

The operationalisation of each indicator also varied substantially, and the details can be found in supplementary material 4. Education was measured primarily as a categorical variable (e.g., level of qualification, n = 25), a binary variable (e.g., having a degree or not, n = 12), a continuous variable (e.g., years of schooling, n = 9), or other approaches as proxy measure for education (e.g., number of books in home, n=2; age at leaving school, n = 1).

The indicators used to measure income/wealth were total household wealth (n = 15), perceived income/wealth (n = 14), household income (n = 8), expenditure measures (n = 4), asset measures (n = 2), individual income (n = 2), pension presence (n = 2), and other economic indicators (n = 3).

Occupational status was measured through several approaches. Occupational level classifications were the most frequently used (n = 13), followed by employment status (n = 3), other occupational measures (n = 3), agricultural versus non-agricultural employment categories (n = 3), and manual versus non-manual work distinctions (n = 1). Within occupational level classifications, three-category systems were most common (n = 9),

Furthermore, to capture a broader socio-economic context, 17 studies examined childhood family background (such as parental education or occupation), while 5 studies incorporated housing conditions and crowding indicators into their analytical frameworks, and 2 studies included neighbourhood-level indicators.

### 3.3 Association between SEP Indicators and Healthy Ageing

The detailed findings from each study are presented in Table 2 and 3. A narrative synthesis of the main findings is presented below.

#### 3.3.1 Education

43 studies examined the relationship between education and healthy ageing. Of these, 39 (90.7%) found a protective effect of higher education on healthy ageing. The remaining 4 studies showed mixed results: 1 with partial support, 1 with no association among men, 1 with no association after adjusting for midlife characteristics such as occupation, and 1 with associations that varied depending on model specification (significant in some models but not others).

The remaining 4 studies showed more complex patterns: 1 demonstrated that the effect of education was mediated through midlife characteristics rather than having a direct effect. 1 found age-dependent effects (significant at age 77, but not at 90). 1 reported gender-specific effects (there was no association among men). 1 found that the protective effects of education varied by healthy ageing model. These effects were significant for the Rowe and Khan model and the psychosocial model, but not for the biomedical model or the complete model.

Similarly, regarding the effect on the rate of change in healthy ageing indicators, the evidence showed consistent protective effects. Of the 18 studies examining this relationship, 16 studies found that a higher level of education was significantly associated with a slower rate of health deterioration (e.g., McLaughlin et al., β = 0.022, p = 0.002). The remaining 2 studies showed small or non-significant effects on change rates (e.g., Wu et al., 2020; β = 0.04, 95% CI: 0.00, 0.09), though education still influenced baseline levels of health (e.g., Wu et al., 2020; β = 10.54, 95% CI: 10.31, 10.77).

#### 3.3.2 Income and Wealth

Among the 36 studies that examined the relationship between income or wealth indicators and healthy ageing, 31 (86.1%) reported a protective effect. Among the remaining 5 studies, 2 studies found no association between income/wealth indicators and healthy ageing (for example, Nurrika et al., 2020; OR = 0.88, 95% CI: 0.60, 1.30), and 3 studies reported partial associations (with protective effects observed only in specific subgroups, countries, or analytical models).

Regarding the effect on healthy ageing, most evidence indicated that higher income or wealth was mainly associated with higher initial levels of health. However, the evidence for its impact on the rate of change was inconsistent: for example, Stolz et al. (2017) reported a convergence effect, where the gap narrowed with age (unstandardized regression coefficient [γ] = 0.003, 95% CI: 0.001, 0.005), while Wu et al. (2020) found no significant effect.

#### 3.3.3 Occupation

22 studies specifically examined the relationship between occupation and healthy ageing, of which 14 (63.6%) found a significant association. The remaining 8 studies (36.4%) showed different results, including null associations, gender- or model-specific effects, associations with varying strength and significance across countries, and in 1 study, occupational effects became non-significant after adjusting for intellectual abilities.

#### 3.3.4 Other SEP Indicators

Beyond the traditional indicators of education, income, and occupation, a few studies reported associations between healthy ageing and other socio-economic factors, such as housing conditions (e.g., Whitley et al., 2018; The Slope Index of Inequality [SII] = 2.66, 95% CI: 2.36, 2.96), and neighbourhood resources (e.g., Liu et al., 2023; β = −0.053, p<0.001). The long-term effects of childhood family background are analysed in Section 3.4.1.

### 3.4 Patterns of Life Course SEP Effects on Healthy Ageing

#### 3.4.1 Long-term Effects of Childhood SEP

Seventeen studies examined childhood SEP using parental occupation (n=13), parental education (n=10), or childhood economic circumstances (n=4) as indicators. Most (n=13) found that disadvantaged childhood circumstances predicted poorer baseline health levels in later life.

For example, the study by Cheval showed that having fewer books in the childhood home was significantly associated with poorer verbal fluency (β = −1.84, p < 0.001) and lower muscle strength (β = −0.93, p < 0.001) in men during adulthood. In Mexico, Grimard and colleagues found that men with poorer childhood socio-economic conditions (as measured by the CSES index) showed 16.2 percentage points lower probability of being healthy in later life for each unit increase in the index (marginal effect = −0.162).

#### 3.4.2 Cumulative Effects

Five studies examined the cumulative effect of life-course socio-economic disadvantage on healthy ageing. All studies found that cumulative disadvantage was associated with worse health outcomes, with effect sizes typically larger than those for exposure at any single time point.

For example, the cumulative inequality index constructed by Whitley had an effect size (absolute difference in SA score [SII] = 4.38, 95% CI: 3.98, 4.78) that exceeded that of any single indicator. Using latent class analysis, Harber-Aschan identified a ‘lifelong low SES’ group; this group not only had the lowest initial health level (β = −0.45, 95% CI: −0.62, −0.29) but also the fastest rate of health decline (β for time interaction = −0.08, 95% CI: −0.11, −0.06). The study by Foverskov also found that, compared to individuals who never experienced financial hardship, those who experienced four or more years of hardship had significantly poorer mid-life physical function (e.g., grip strength) (*b* = −1.22 kg, 95% CI: −2.38, −0.07).

#### 3.4.3 Social Mobility and Health Trajectories

Three studies analysed the impact of social mobility on healthy ageing. All found that upward mobility was associated with better health outcomes and downward mobility was associated with worse healthy ageing.

For example, Huang found that upward social mobility groups had higher intrinsic capacity scores compared to persistently low SEP groups (β = 1.57, 95% CI: 1.42, 1.73), while downward social mobility groups had lower scores compared to stable high SEP groups (β = −0.63, 95% CI: −0.90, −0.36). Payne reported that men on a ‘high-to-low’ mobility trajectory had lower disability-free life expectancy at age 45 (24.35 years) compared with men on a stable ‘high-to-high’ trajectory (30.29 years).

#### 3.4.4 Age-related Patterns of Health Inequality

23 studies identified the dynamic evolution of health inequality with age. These studies employed two primary analytical approaches: 11 studies utilized variable-centred methods, such as time-interaction terms or growth curve models, to directly quantify the rate of health change. The remaining 12 studies used person-centred approaches, identifying distinct groups of individuals with qualitatively different health trajectories (e.g., “stable,” “slow decline,” “rapid decline”) and analysing predictors of group membership.

Across these studies, three patterns of evolution were observed. Most studies (18/23) supported a divergence pattern, with health disparities widening over time. The divergence pattern was most consistently found when education was the SEP indicator and for outcomes of cognitive or overall health. For example, García-Esquinas reported that lower educational attainment was linked to a more rapid accumulation of health deficits (slope β=0.84 for lowest education vs. β=0.60 for highest). Harber-Aschan also found that the low SEP group had a significantly steeper slope of health decline (interaction β = −0.08). This pattern of widening inequality was domain-specific, with Cheng reporting that socioeconomic differences in cognitive health became larger over time, while differences in physical health remained stable (Incidence Rate Ratio [IRR] = 1.04, 95% CI: 1.03, 1.05).

A convergence pattern was observed in 2 studies, where health disparities narrowed with increasing age, primarily for physical health outcomes when using income as the SEP indicator. For example, the study by Stolz reported that income-related gaps in frailty converged, with a faster growth rate in frailty among higher-income groups (Third vs. first income quartile: γ = 0.004, 95% CI: 0.001, 0.007). Cheng also found that the association of income with multimorbidity weakened in older age (IRR = 1.12, 95% CI: 1.10, 1.14).

The third pattern, persistent inequality, was supported by 3 studies showing that health disparities established earlier in life remained relatively stable in older age. For example, Cheval and Wu found that SEP primarily influenced baseline health levels, with minimal impact on the rate of change over time.

### 3.5 Contextual Effects and Statistical Associations

#### 3.5.1 Cross-regional and Contextual Variations

The studies included in this review spanned over 20 countries and regions, with 25 studies from high-income countries and 22 from or including samples from low- and middle-income countries (LMICs).

A positive association between education and healthy ageing was consistently reported across different regions, with 39 of the 43 studies examining this indicator finding a significant effect. However, the role and measurement of other SEP indicators demonstrated variations by geographical and social context.

The analysis of social stratification factors revealed distinct priorities for different regions. Among the 12 studies from mainland China and Taiwan, 6 reported the hukou system (household registration system affecting access to services) or urban-rural residence as a significant variable in healthy ageing, with urban residence and non-agricultural hukou status generally associated with better health outcomes (Li, 2022; Payne, 2022; Si, 2023; Chang, 2023; Chen 2024; Wang, 2024). Among 10 studies from North American populations, 5 included race/ethnicity as a core variable or stratification factor in their analyses, typically showing persistent health disparities with minority populations experiencing higher risks of poor health trajectories compared to whites. However, some studies found that higher education may mitigate racial disparities, particularly for African Americans. (Xu, 2014; Pruchno, 2015; de la Fuente, 2018; McLaughlin, 2020; Lu, 2021).

The relative importance of different economic indicators also varied. Cross-national comparative studies reported that wealth had stronger protective effects than income in the USA (β = −0.033 vs. −0.014), whereas income was more protective than wealth in China (β = −0.032 vs. −0.007) (Lu et al. 2021).

#### 3.5.2 Moderation, Mediation, and Bidirectional Associations

Five studies reported significant moderation effects by gender. White found that the effect of occupation on healthy ageing was significant only in men (OR = 2.60, 95% CI: 1.14, 6.05), but not in women (OR = 1.67, 95% CI: 0.93, 3.04). Similarly, Cosco found education significantly predicted higher functional trajectories in women (OR = 1.50, 95% CI: 1.11, 2.03) but not in men (OR = 1.31, 95% CI: 0.90, 1.92).

Six studies used methods such as structural equation modelling or causal mediation analysis to examine mediating pathways. For example, Wu reported that adult SEP (education and wealth) mediated between 21% and 78% of the association between childhood SEP and healthy ageing scores across different populations.

Beyond moderation and mediation, one study examined bidirectional associations between SEP and health. Significant bidirectional effects were found in men, with standardised path coefficients of β = 0.055 (95% CI: 0.047, 0.064) from baseline wealth to change in healthy ageing outcomes (e.g., grip strength) and β = 0.045 (95% CI: 0.036, 0.055) for the healthy ageing to wealth pathway (Ahrenfeldt et al. 2021).

## 4. Discussion

This systematic review represents the first comprehensive synthesis specifically examining longitudinal studies on the relationship between SEP and healthy ageing, whilst analysing the dynamic associations between SEP across the life course and healthy ageing. The review included 47 studies spanning over 20 countries and regions, comprising 27 studies (57.4%) from high-income countries and 20 (42.6%) from low- and middle-income countries. Risk of bias assessment indicated high overall evidence quality, with only five studies rated as high risk. Despite substantial methodological heterogeneity across studies in the definitions of healthy ageing and the measurement and operationalisation of SEP indicators, the overall findings remained consistent. Education emerged as the most consistently protective indicator, with 90.7% of studies reporting positive associations, followed by income/wealth (86.1%), while occupation-related indicators demonstrated comparatively lower support (63.6%).

### 4.1 SEP and healthy ageing

#### 4.1.1 Education and healthy ageing

The protective role of education may operate through two distinct pathways: initial advantage and sustained protection. Education, established early in life, enhances health knowledge and behavioural management capabilities while improving employment opportunities and income levels (Leoni, 2025). This establishes stronger foundations for living healthier lives for individuals. Furthermore, the cognitive reserve developed through education gives the brain redundant capacity to cope with pathological changes (Stern, 2012). Individuals with more years of schooling may therefore demonstrate greater adaptability and problem-solving abilities when facing health challenges.

Four studies suggest that although educational advantages are generally robust, their expression may vary depending on intervening life course factors (such as midlife status), demographic characteristics, and how healthy ageing is conceptualized and measured.

The measurement of education is subject to variation across studies and between countries, thereby impacting the comparability of results. Nevertheless, education as a health protective factor has been validated across diverse institutional and cultural contexts, demonstrating considerable universality.

Education should therefore be regarded as a lifelong health investment, with the WHO Decade of healthy ageing: baseline report emphasising the importance of lifelong learning opportunities for maintaining cognitive and functional capacity. In view of the cross-cultural consistency of educational effects and their dual protective mechanisms, the expansion of educational opportunities and the enhancement of educational quality should be considered as significant policy directions for the promotion of healthy ageing.

#### 4.1.2 Income, wealth and healthy ageing

Economic resources exhibit complex patterns of influence on healthy ageing. Of the 36 studies reviewed, 31 (86.1%) reported positive associations.

Economic resources can improve material living conditions directly by enhancing purchasing power and accessibility to resources, including nutrition, housing and healthcare (Finkelstein et al., 2022; French et al., 2019; Galobardes et al., 2007). However, income and wealth are not identical concepts. Income primarily reflects current consumption capacity, determining whether individuals can afford daily health expenditures. In contrast, wealth represents asset accumulation and may provide long-term financial security for health needs, although this may depend on asset liquidity and the strength of social security systems (Machado et al., 2025; Tao et al., 2025).

This distinction is particularly significant in later life. Despite having relatively fixed or declining incomes, many older adults may have accumulated considerable wealth in the form of property, savings accounts and other financial assets, resulting in a ‘low income, high wealth’ economic profile. In such circumstances, wealth may be a more accurate reflection of actual economic capacity (Gugushvili and Wiborg, 2025). Furthermore, subjective economic perceptions (such as perceptions of income adequacy) exert an independent influence on healthy ageing. Even at equivalent objective income levels, these perceptions may promote health through psychosocial pathways including stress reduction and enhanced sense of control (Feng et al., 2015). The importance of economic indicators also varies according to societal environment. Research found that wealth inequalities were more pronounced than income inequalities in healthy ageing in the United States, while income was a more influential predictor than wealth in China. Such differences may stem from variations in healthcare payment systems: high out-of-pocket medical costs in the US make accumulated wealth crucial for healthcare access (Himmelstein et al., 2009), whereas health insurance coverage in China makes current income more important (Zhang et al., 2015).

Nevertheless, five studies found no significant associations with income/wealth indicators. However, these apparent null findings may still support economic resources’ relevance through different pathways: the independent effect of wealth became non-significant after controlling for the highly correlated variable of education in Mexico; in Canada, subjective income measures mediated the effect of education on healthy ageing for men, but this pathway was not observed for women; higher expenditure showed no association in Indonesia, which researchers suggested could be due to higher-income groups adopting unhealthy dietary and lifestyle habits that offset their economic advantages; and cross-national patterns emerged where income showed no associations in UK/Japan while wealth showed none in China, likely reflecting welfare system differences. These patterns suggest that, unlike education which showed consistent protective effects (though educational attainment itself partly reflects economic resources through access barriers), economic resources operate through more context-dependent pathways in their associations with healthy ageing.

Overall, economic support policies for older adults should address not only routine income enhancement, such as pensions, but also asset protection, wealth accumulation and improving economic security. Income subsidies alone may be insufficient to improve health outcomes in older populations. Additionally, universal health coverage policies may help neutralise income and wealth-based health inequalities by reducing financial barriers to healthcare access.

#### 4.1.3 Occupation and healthy ageing

The protective effects of occupation on healthy ageing are relatively inconsistent. Of the 22 studies, 14 (63.6%) reported protective effects, but the results demonstrated considerable heterogeneity. This reflects the structural complexity of occupation as an SEP indicator, as well as the wide variety of measurement approaches used.

The concept of occupation encompasses multiple attributes, including economic returns, social prestige, working conditions and employment stability (Fujishiro et al., 2010). Different measurement approaches in studies generate distinct predictive pathways. For example, classifications based on occupational prestige primarily capture the effects of social status, operating through social resources and lifestyle factors (Christ et al., 2012). In contrast, classifications based on physical labour intensity more closely reflect health risk exposure, with a direct link to physical wear and occupational disease risk (Eyles et al., 2019).

There are significant gender differences in occupational effects. Traditional occupational classification systems are often based on the experiences of men in the labour market and may not accurately reflect the actual SEP of women. Research suggests that occupational effects are more pronounced among men, which may be related to greater career continuity and a stronger dependence of identity on occupational achievement (Schellenberg et al., 2016). In contrast, women’s employment pathways are often shaped by disproportionate caregiving responsibilities and social expectations around domestic labour, roles that remain unequally distributed due to sexism. For many women, caregiving can become a central aspect of life course experience (Ishizuka and Musick, 2021; Mussida and Patimo, 2021), yet this is frequently under- or non-represented in conventional SEP research, which often prioritises paid work.

The applicability of occupational classifications also faces challenges against the backdrop of rapid social transition. Socio-economic transformation often reshapes occupational prestige systems, with some previously high-status occupations experiencing status decline during transition periods, while the social status evaluation of emerging occupations remains unstable (Newlands and Lutz, 2024; Ulfsdotter Eriksson et al., 2022). Furthermore, characteristics such as high proportions of informal employment and substantial occupational mobility mean that traditional occupational classifications may fail to accurately reflect current social status structures (Lin and Hung, 2022).

Overall, the influence of occupation on healthy ageing is highly context-dependent. Its relationship with healthy ageing may depend not only upon the classification dimensions employed but is also constrained by the social institutional background and economic development stage of the country/region concerned. Researchers should therefore select classification approaches that best reflect the actual social position of target populations according to specific social contexts and research objectives.

More broadly, this context-dependency extends beyond occupation to other SEP indicators. For example, Chinese studies universally focus on the hukou registration system, while North American studies emphasise racial and ethnic factors, suggesting that standard SEP frameworks may have limitations when applied across cultures (Howe et al., 2012). Effective health equity research requires identification of stratification dimensions with actual influence within local social structures.

### 4.2 SEP and healthy ageing from a life course perspective

Evidence from life courses indicates that current SEP cannot fully explain differences in healthy ageing; early experiences and cross-stage cumulative effects may be equally important (Huang et al., 2025; Payne and Xu, 2022; Wagg et al., 2021).

The long-term influence of childhood SEP is particularly significant. Of the 17 studies, 13 found that disadvantage in childhood SEP predicted poorer health outcomes in later life. This effect remained consistent even when controlling for adult SEP. This finding supports the sensitive period hypothesis, which suggests that the origins of health inequalities may be established early in life (Cable, 2014; Kuh et al., 2003). The risk chain model helps to explain this transmission mechanism: low SEP in childhood may initiate a series of subsequent risks (Ben-Shlomo and Kuh, 2002; Bowen and González, 2010). Poor childhood living environments and family economic pressures not only affect children’s nutrition, development, and health status but also limit educational opportunities, thereby influencing adult occupational choices, income levels, and social position (Lynch et al., 1997). This creates a clear pathway from early disadvantage to later poor health. The long-term influence of childhood SEP therefore reflects the path-dependent relationship between early social positioning and subsequent life trajectories.

Five studies supported the notion of cumulative effects, showing that multi-stage SEP disadvantage may have further detrimental effect. Furthermore, individuals with lifelong low SEP trajectories not only had lower health baselines, but also experienced a more rapid decline, exhibiting a “double burden” pattern. These findings align with cumulative disadvantage theory (Dannefer, 2003; O’Rand, 1996). The accumulation of adverse conditions throughout the life course continuously depletes individuals’ adaptive reserves, including material resources, social support, and psychological resilience, thereby undermining their capacity to cope with ageing challenges (Barboza Solís et al., 2015; Liu et al., 2023b). When physiological challenges associated with ageing emerge, individuals with fewer reserves may appear more vulnerable.

Three studies on social mobility found that changes in SEP significantly influence healthy ageing. Upward mobility is associated with better health improvements, though psychological adaptation challenges may limit these gains (Islam and Jaffee, 2024). Downward mobility produces more rapid health deterioration, consistent with asymmetric responses to health deterioration versus improvement (Binder and Coad, 2013; Islam and Jaffee, 2024; Miller et al., 2020). However, reverse causation should be considered, whereby poor health may contribute to downward mobility.

One study identified a group with “mixed SEP trajectories” featuring economic instability, which had poorer health than those with persistently low SEP and experienced the fastest health decline. This suggests SEP instability itself may be a more important health risk factor than absolute SEP levels, as continuous economic fluctuations create chronic uncertainty and sustained psychosocial stress (Frasquilho et al., 2015; Ryu and Fan, 2023).

Health inequality patterns with age showed three trends among 23 studies: 18 supported widening disparities, consistent with cumulative disadvantage theory (Dannefer, 2003; Ferraro and Shippee, 2009; O’Rand, 1996), while a minority found convergence or persistent patterns formed in early life.

The widening trend was most evident for education and cognitive function, where cognitive reserve theory suggests that educational resources provide protection against age-related decline (Stern, 2012). Physical function showed different patterns, with some income-related inequalities converging in later life due to universal functional decline and selective survival of healthier individuals (Hoffmann, 2011; McMunn et al., 2009).

These life course patterns are evident not only at the individual level, but also at the population level, where they demonstrate intergenerational differences. Research has found that the protective effects of wealth on health are stronger among younger age groups. This suggests that high-wealth groups primarily benefit from social progress and medical advances, which leads to a further widening of health inequalities between age groups (Link and Phelan, 1995).

### 4.3 Complex association patterns

Beyond the direct effects of individual SEP indicators, this review reveals more complex relationships between SEP and healthy ageing.

Mediation analyses reveal the diversity of pathways through which SEP influences health. Adult SEP also demonstrates significant mediating effects in the pathways through which childhood SEP influences health, with explanatory proportions ranging from 21% to 78%.

This suggests that midlife may represent a critical yet underexplored period for healthy ageing interventions. While childhood and late-life factors have received considerable attention, the middle years (when many health trajectories become established) may remain relatively understudied in healthy ageing research. The plasticity of health outcomes during midlife may present important opportunities for interventions that could alter long-term ageing trajectories, warranting greater research attention.

Research on bidirectional relationships between SEP and health is scarce, with only one of the 47 studies addressing this issue. This study revealed that poor health makes it harder to maintain a favourable SEP, while a decline in SEP further damages health, creating a cycle of disadvantage (Álvaro et al., 2023; Xiong and Qi, 2024). This bidirectionality complicates causal identification in longitudinal studies and requires more research attention.

### 4.4 Strengths and Limitations

This review has several strengths. It is the first systematic review to focus exclusively on the relationships between SEP and healthy ageing in longitudinal studies. It incorporates 47 studies, involving over 20 countries, with follow-up periods of up to 23 years. We systematically examined heterogeneity in SEP and healthy ageing measurements, distinguished between single-stage and multi-stage SEP indicators, presented the characteristics of the research in evidence tables, and explored complex associations from a life-course perspective. This provides an empirical foundation for understanding the long-term mechanisms of health inequality.

This review also has some limitations. First, around half of the studies (24/47) may be subject to the ‘Table 2 fallacy’, whereby the influence of control variables is incorrectly interpreted as independent causal effects. Second, the heterogeneity of SEP and healthy ageing measurements restricts the ability to compare effect sizes and precludes quantitative meta-analysis. Third, our focus on participants aged 50 years and above may have limited insights into earlier life course transitions. Fourth, overlapping use of large cohort datasets may lead to concentrated evidence sources, requiring cautious interpretation of conclusion robustness. Fifth, studies from low-income countries in Africa and Latin America are relatively scarce. Finally, search strategies limited to specific databases and English-language publications, may have introduced selection bias.

Future research should therefore focus on the following areas: first, employing robust methods informed by causal inference to clarify bidirectional relationships between SEP and healthy ageing. Second, core measurement indicator sets with cross-cultural adaptation could be established to improve study comparability. This will include domain-specific assessments to examine the differential effects of SEP across health domains. Furthermore, research should examine the earlier origins of health inequalities by investigating SEP-health relationships from midlife or even younger ages, as studies beginning at age 50 and above may miss key socioeconomic transitions that shape healthy ageing trajectories. Future work should also explore health effects of social mobility and independent mechanisms of SEP instability.

Finally, high-quality longitudinal studies from underserved regions such as Africa and Latin America are needed to verify the universality of findings and provide further understanding of the contextual factors that may impact SEP-healthy ageing relationships.

## 5. Conclusion

This systematic review confirms significant associations between SEP and healthy ageing. Education has the most consistent protective effect, while the influence of income and wealth is positive but complex. Evidence regarding occupation and healthy ageing is relatively inconsistent. Life course evidence suggests that childhood socio-economic circumstances have a lasting impact on healthy ageing, with health inequalities tending to widen in later life. However, evidence regarding dynamic changes in midlife SEP and the health effects of social mobility remains limited.

Policy interventions for healthy ageing inequalities require targeted strategies. First, given the cross-culturally consistent protective effects of education, particular emphasis should be placed on early educational investment, especially early life interventions for educationally disadvantaged groups, as well as lifelong learning opportunities throughout the life course. Second, diversified economic support models should be adopted to address asset protection, wealth accumulation mechanisms, and enhancement of economic security amongst older adults. Finally, differentiated intervention strategies should be implemented across different life stages, from childhood socio-economic environment improvements to diversified support services in later life.

Although this review incorporates studies from over 20 countries, evidence from certain social, institutional, and cultural contexts remains insufficient. Further research is needed to explore the mechanisms of action between SEP and healthy ageing in more detail, particularly about the optimal timing and approaches for interventions within different sociocultural contexts.

## Supporting information

Supplementary material 1. checklist

Supplementary material 2. criteria and search strategy

Supplementary material 3. quality assessment

Supplementary material 4. heterogeneity

## Author contributions

All authors contributed to the study conception and design. YY conducted the searches, screening, extraction, data charting, and analysis. CL undertook screening and risk of bias assessment stages. DMA acted as the third reviewer. YY drafted the initial manuscript. DMA, KC and MP critically reviewed and edited the draft. All authors read and approved the final manuscript.

## Funding

YY is supported by the King’s-China Scholarship Council PhD Scholarship Programme. DMA is supported by the Wellcome Trust [grant number 304283/Z/23/Z] and by the Economic and Social Research Council (ESRC) Centre for Society and Mental Health at King’s College London [grant number ES/S012567/1].

## Declaration of Competing Interest

All authors declare no competing interests. The funder had no role in study design, data collection and analysis, decision to publish, or preparation of the manuscript.

## Data availability

The datasets used and/or analysed during the current study are available from the corresponding author on reasonable request.

## Notes

### Competing Interest Statement

The authors have declared no competing interest.

### Summary of Updates

An error in Table 1 has been corrected, and the format of the references has been amended.

