## Supplementary material 2. criteria and search strategy for "Life course socio-economic position and healthy ageing: A systematic review of longitudinal studies"

**Inclusion Criteria**

Our systematic review will include studies that:

1. were longitudinal, observational, retrospective, and prospective studies;

2. were conducted in middle-aged or older adults (studies that include multiple populations with various age groups will only be included if the mean age of the sample is 50 or over at baseline);

3. incorporated a combined measure of healthy ageing or report on at least two domains separately (i.e., physical functioning, cognitive functioning, mental health, absence of disease, participation in activities, personal perception, and healthy survival);

4. measured socioeconomic factors at a minimum of two time points and measured healthy ageing at a minimum of one time point for research question one;

5. measured socioeconomic factors at a minimum of one time point and measured healthy ageing at multiple time points for research question two; and

6. were published articles or theses from English-language peer-reviewed journals.

**Exclusion criteria**

Our systematic review will exclude studies that:

1. were intervention studies/trials, case-control studies, cross-sectional studies, and ecological studies;

2. include participants from hospitalised or disease specific cohorts;

3. include participants with various age groups, and the mean age of the sample is less than 50;

3. do not investigate socioeconomic factors;

4. do not report any association between socioeconomic factors and healthy ageing;

5. included a single measurement of both healthy ageing and socioeconomic factors at one time point;

6. exclusively reported a single domain of healthy ageing, such as cognitive or physical functioning; and

7. were articles or theses published in non-English language journals.

**Search strategy**

#1 – identifying life course studies that included more than one measure of risk factor over life.

#2 – identifying studies with socio-economic variables.

#3 – identifying studies with the main outcome (any definition related to ‘healthy ageing’)

#4 – #1 AND #2 AND #3

#5 – Remove duplicates from 4

Table 1. Search terms used for Embase, Psychinfo (OVID interface)

|  | Search terms |
| --- | --- |
| #1 | ("lifecourse" OR "life course" OR "life-course" OR "life-stage*" OR "life-trajector*" OR "life-path*" OR "life-transition*" OR "life-chang*" OR "lifelong" OR "life-long" OR "life long" OR "grow-up" OR "growing-up" OR "grown-up*" OR "grownup*" OR "life-experience*" OR "turning-point*" OR "long term" OR "long-term*" OR "critical period" OR "sensitive period" OR "social mobility" OR "traject*" OR ((upward* OR downward* OR mobility) AND (social OR socioeconomic OR socio-economic OR economic* OR education* OR occupation*)) OR "early life" OR "later-life" OR "later-in-life" OR "child*" OR "adolescen*" OR "cumulat*" OR "accumulat*") |
| #2 | ("ses" OR "socioeconomic*" OR "socio-economic*" OR "social class" OR "social position" OR "social status" OR "social conditions" OR "social capital" OR ((social OR occupational OR economic OR educational) AND (inequalit* OR disparit* OR inequit*)) OR "income" OR "occupation*" OR "poverty" OR "remuneration" OR "salaries and fringe benefits" OR "education" OR "educational status" OR "employ*" OR "unemploy*" OR "impoverished" OR "affluen*" OR "advantage*" OR "disadvantage*" OR "depriv*" OR "inequalit*" OR "wealth*" OR "earning*" OR "wage*") |
| #3 | ("health* ag?ing" OR "successful ag?ing" OR "active ag?ing" OR "meaningful ag?ing" OR "effective ag?ing" OR "robust ag?ing" OR "unimpaired ag?ing" OR "positive ag?ing" OR "productive ag?ing" OR "ag?ing well" OR "optim* ag?ing" OR "healthy longevity" OR "exceptional survival" OR "healthy elder*" OR "healthy older*") |

Table 2. Search terms used for Medline (PubMed interface)

|  | Search terms |
| --- | --- |
| #1 | (("lifecourse") OR ("life course") OR ("life-course") OR ("life-stage*") OR ("life-trajector*") OR ("life-path*") OR ("life-transition*") OR ("life-chang*") OR ("lifelong") OR ("life-long") OR ("life long") OR ("grow-up") OR ("growing-up") OR ("grown-up*") OR ("grownup*") OR ("life-experience*") OR ("turning-point*") OR ("long term") OR ("long-term*") OR ("critical period") OR ("sensitive period") OR ("social mobility") OR ("traject*") OR (("upward*" OR "downward*" OR "mobility") AND ("social" OR "socioeconomic" OR "socio-economic" OR "economic*" OR "education*" OR "occupation*")) OR ("early life") OR ("later-life") OR ("later-in-life") OR ("child*") OR ("adolescen*") OR ("cumulat*") OR ("accumulat*") OR "Life Course Perspective"[Mesh Terms] OR "Life Change Events"[Mesh Terms])) |
| #2 | (("ses") OR ("socioeconomic*") OR ("socio-economic*") OR ("social class") OR ("social position") OR ("social status") OR ("social conditions") OR ("social capital") OR (("social" OR "occupational" OR "economic" OR "educational") AND ("inequalit*" OR "disparit*" OR "inequit*")) OR ("income") OR ("occupation*") OR ("poverty") OR ("remuneration") OR ("salaries and fringe benefits") OR ("education") OR ("educational status") OR ("employ*") OR ("unemploy*") OR ("impoverished") OR ("affluen*") OR ("advantage*") OR ("disadvantage*") OR ("depriv*") OR ("inequalit*") OR ("wealth*") OR ("earning*") OR ("wage*") OR "Socioeconomic Factors"[MeSH Terms])) |
| #3 | (("health* ageing" OR "health* aging" OR "successful ageing" OR "successful aging" OR "active ageing" OR "active aging" OR "meaningful ageing" OR "meaningful aging" OR "effective aging" OR "robust aging" OR "unimpaired aging" OR "positive ageing" OR "positive aging" OR "productive ageing" OR "productive aging" OR "ageing well" OR "aging well" OR "healthy elder*" OR "healthy older*" OR "healthy longevity" OR "exceptional survival" OR “Healthy Aging"[MeSH Terms])) |

Table 3. Search terms used for the Web of Science (All databases)

|  | Search terms |
| --- | --- |
| #1 | TS=("lifecourse" OR "life course" OR "life-course" OR "life-stage*" OR "life-trajector*" OR "life-path*" OR "life-transition*" OR "life-chang*" OR "lifelong" OR "life-long" OR "life long" OR "grow-up" OR "growing-up" OR "grown-up*" OR "grownup*" OR "life-experience*" OR "turning-point*" OR "long term" OR "long-term*" OR "critical period" OR "sensitive period" OR "social mobility" OR "traject*" OR ((upward* OR downward* OR mobility) AND (social OR socioeconomic OR socio-economic OR economic* OR education* OR occupation*)) OR "early life" OR "later-life" OR "later-in-life" OR "child*" OR "adolescen*" OR "cumulat*" OR "accumulat*") |
| #2 | TS=("ses" OR "socioeconomic*" OR "socio-economic*" OR "social class" OR "social position" OR "social status" OR "social conditions" OR "social capital" OR (("social" OR "occupational" OR "economic" OR "educational") AND ("inequalit*" OR "disparit*" OR "inequit*")) OR "income" OR "occupation*" OR "poverty" OR "remuneration" OR "salaries and fringe benefits" OR "education" OR "educational status" OR "employ*" OR "unemploy*" OR "impoverished" OR "affluen*" OR "advantage*" OR "disadvantage*" OR "depriv*" OR "inequalit*" OR "wealth*" OR "earning*" OR "wage*") |
| #3 | TS=("health* ageing" OR "health* aging" OR "successful ageing" OR "successful aging" OR "active ageing" OR "active aging" OR "meaningful ageing" OR "meaningful aging" OR "effective ageing" OR "effective aging" OR "robust ageing" OR "robust aging" OR "unimpaired ageing" OR "unimpaired aging" OR "positive ageing" OR "positive aging" OR "productive ageing" OR "productive aging" OR "ageing well" OR "aging well" OR "optim* ageing" OR "optim* aging" OR "healthy elder*" OR "healthy older*" OR "healthy longevity" OR "exceptional survival") |
