## Supplementary material 3. quality assessment for "Life course socio-economic position and healthy ageing: A systematic review of longitudinal studies"

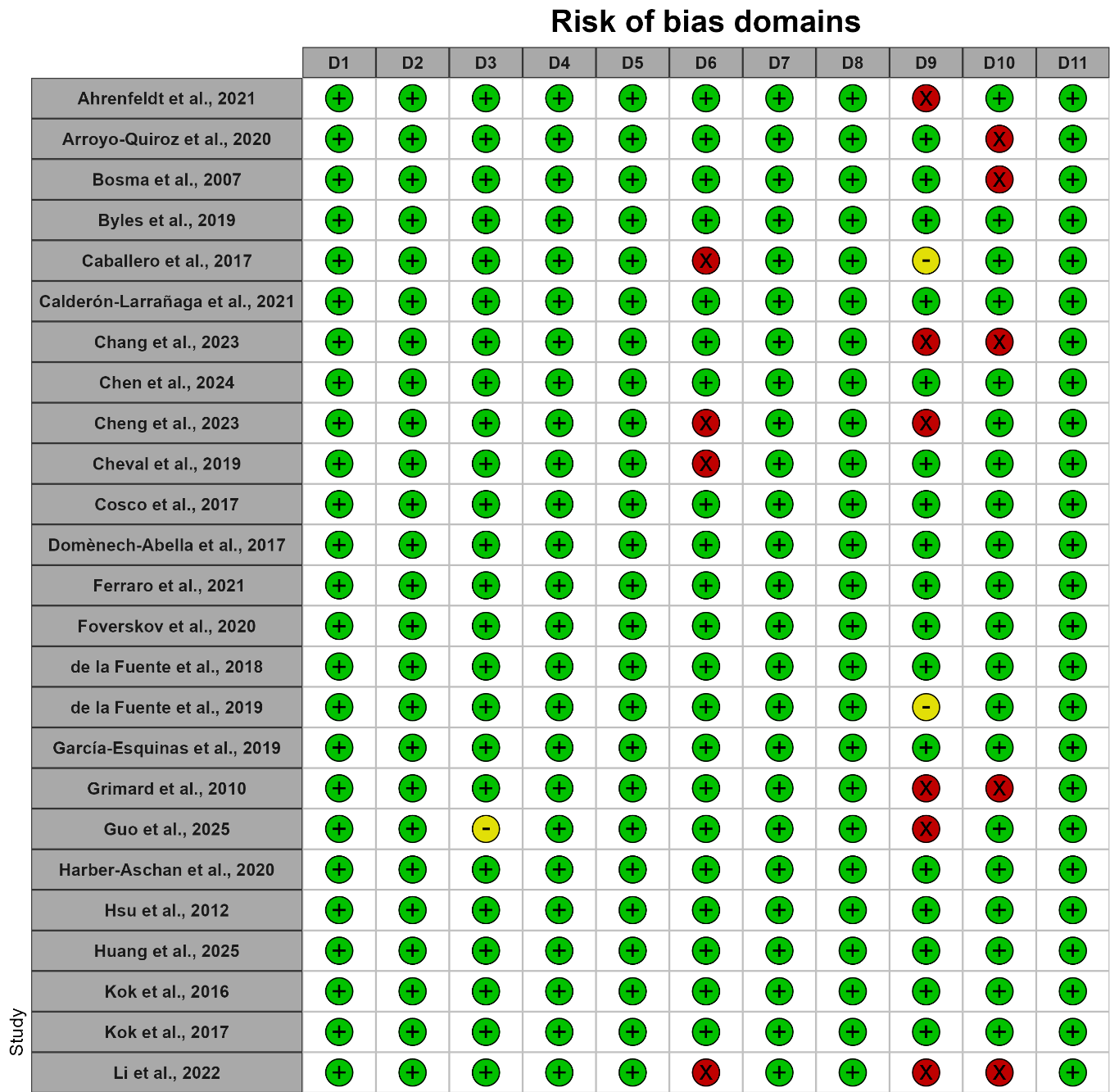

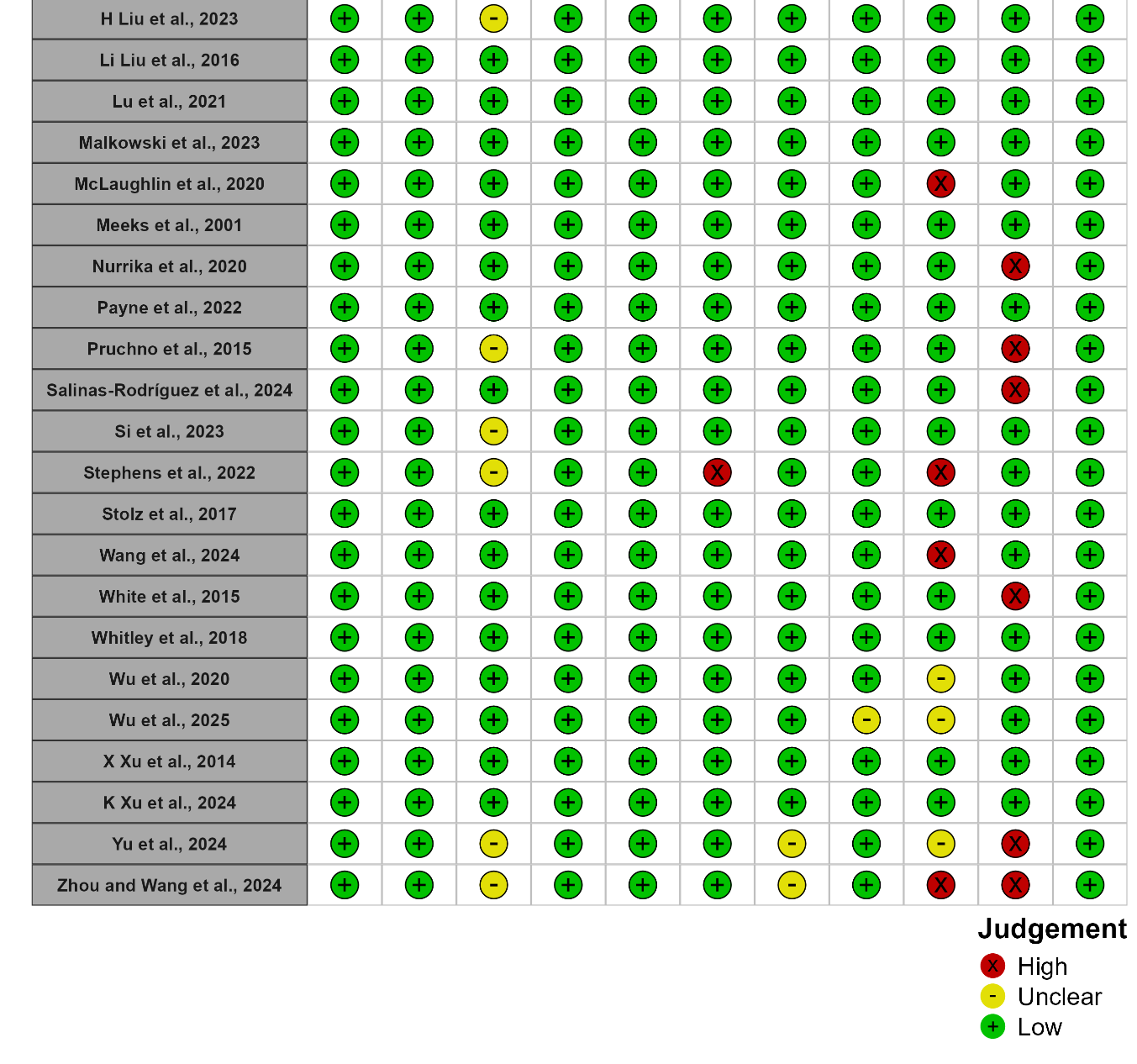

Figure S1: Risk of bias assessment for included studies using the Joanna Brigg’s Institute critical appraisal checklist for cohort studies.

Domains:

D1: Were the two groups similar and recruited from the same population?

D2: Were the exposures measured similarly to assign people to both exposed and unexposed groups?

D3: Was the exposure measured in a valid and reliable way?

D4: Were confounding factors identified?

D5: Were strategies to deal with confounding factors stated?

D6: Were the groups/participants free of the outcome at the start of the study (or at the moment of exposure)?

D7: Were the outcomes measured in a valid and reliable way?

D8: Was the follow up time reported and sufficient to be long enough for outcomes to occur?

D9: Was follow up complete, and if not, were the reasons to loss to follow up described and explored?

D10: Were strategies to address incomplete follow up utilized?

D11: Was appropriate statistical analysis used?
