## Supplementary material 4. heterogeneity for "Life course socio-economic position and healthy ageing: A systematic review of longitudinal studies"

**Table 1** Heterogeneity of the healthy ageing included in studies

| **Study** | **Conceptual Framework** | **Physical functioning** | **Cognitive functioning** | **Mental health** | **Absence of disease** | **Participation in activities** | **Personal perception** | **Healthy survival** | **Others** |
| --- | --- | --- | --- | --- | --- | --- | --- | --- | --- |
| Ahrenfeldt et al., 2021 | Biomedical | ✓ | ✓ | ✓ |  |  | ✓ |  |  |
| Arroyo-Quiroz et al., 2020 | Rowe & Kahn | ✓ |  |  | ✓ |  |  | ✓ |  |
| Bosma et al., 2007 | Custom | ✓ | ✓ | ✓ |  |  |  |  |  |
| Byles et al., 2019 | Rowe & Kahn | ✓ |  |  | ✓ |  |  |  |  |
| Caballero et al., 2017 | WHO healthy ageing | ✓ | ✓ | ✓ | ✓ | ✓ | ✓ |  |  |
| Calderón-Larrañaga et al., 2021 | Custom | ✓ | ✓ |  | ✓ |  |  |  |  |
| Chang et al., 2023 | Rowe & Kahn | ✓ | ✓ | ✓ | ✓ | ✓ |  |  |  |
| Chen et al., 2024 | Custom | ✓ | ✓ | ✓ | ✓ | ✓ | ✓ |  |  |
| Cheng et al., 2023 | Biomedical | ✓ | ✓ |  | ✓ |  |  |  |  |
| Cheval et al., 2019 | Custom | ✓ | ✓ | ✓ |  |  | ✓ |  |  |
| Cosco et al., 2017 | Custom | ✓ | ✓ | ✓ |  | ✓ | ✓ |  |  |
| Domènech-Abella et al., 2017 | Custom | ✓ | ✓ | ✓ | ✓ | ✓ | ✓ |  | environmental safety |
| Ferraro et al., 2021 | Custom | ✓ | ✓ |  |  |  |  |  |  |
| Foverskov et al., 2020 | Biomedical | ✓ | ✓ |  |  |  |  |  | Inflammation: C-reactive protein, Interleukin-6, TNF-α |
| de la Fuente et al., 2018 | Custom | ✓ | ✓ |  | ✓ |  | ✓ |  | sensory impairments |
| de la Fuente et al., 2019 | Custom | ✓ | ✓ |  |  |  |  |  | impairments in body functions |
| García-Esquinas et al., 2019 | Deficit Accumulation | ✓ | ✓ |  | ✓ |  | ✓ |  |  |
| Grimard et al., 2010 | Custom | ✓ |  |  |  |  | ✓ |  |  |
| Guo et al., 2025 | Frailty Index | ✓ | ✓ | ✓ | ✓ | ✓ | ✓ |  | ？ |
| Harber-Aschan et al., 2020 | Custom | ✓ | ✓ |  | ✓ |  |  |  |  |
| [Hsu](javascript:;) et al., 2012 | Rowe & Kahn | ✓ |  | ✓ | ✓ | ✓ | ✓ |  |  |
| Huang et al., 2025 | WHO Intrinsic Capacity | ✓ | ✓ | ✓ |  |  |  |  | grip strength, BMI |
| Kok et al., 2016 | Custom | ✓ | ✓ | ✓ |  | ✓ | ✓ |  |  |
| Kok et al., 2017 | Custom | ✓ | ✓ | ✓ |  | ✓ | ✓ |  |  |
| Li et al., 2022 | Custom | ✓ | ✓ |  |  | ✓ |  |  |  |
| H Liu et al., 2023 | Frailty Index | ✓ | ✓ | ✓ | ✓ |  |  |  |  |
| Li Liu et al., 2016 | Custom | ✓ | ✓ | ✓ | ✓ |  | ✓ |  |  |
| Lu et al., 2021 | Custom | ✓ | ✓ | ✓ | ✓ | ✓ | ✓ |  |  |
| Malkowski et al., 2023 | Custom | ✓ | ✓ | ✓ |  | ✓ |  |  | fibrinogen, C-reactive protein |
| McLaughlin et al., 2020 | Custom | ✓ | ✓ |  |  |  |  |  |  |
| Meeks et al., 2001 | Custom | ✓ |  |  |  |  | ✓ |  |  |
| Nurrika et al., 2020 | Custom | ✓ | ✓ |  |  |  | ✓ |  |  |
| Payne et al., 2022 | Health Expectancy | ✓ |  |  |  |  |  | ✓ |  |
| Pruchno et al., 2015 | Custom | ✓ |  |  | ✓ |  | ✓ |  |  |
| Salinas-Rodríguez et al., 2024 | WHO Intrinsic Capacity | ✓ | ✓ | ✓ |  |  |  |  |  |
| Si et al., 2023 | WHO Intrinsic Capacity | ✓ | ✓ | ✓ |  |  | ✓ |  | forced expiratory volume, hemoglobin |
| Stephens et al., 2022 | Custom | ✓ |  | ✓ |  | ✓ |  |  |  |
| Stolz et al., 2017 | Deficit Accumulation | ✓ |  |  | ✓ |  | ✓ |  | body mass index deficit |
| Wang et al., 2024 | WHO healthy ageing | ✓ | ✓ | ✓ |  |  | ✓ |  |  |
| White et al., 2015 | Custom | ✓ | ✓ | ✓ |  | ✓ | ✓ |  |  |
| Whitley et al., 2018 | Rowe & Kahn | ✓ | ✓ |  | ✓ | ✓ |  |  |  |
| Wu et al., 2020 | Custom | ✓ | ✓ |  |  |  |  |  |  |
| Wu et al., 2025 | Custom | ✓ | ✓ |  |  |  |  |  |  |
| X Xu et al., 2014 | Custom | ✓ | ✓ | ✓ |  |  |  |  |  |
| K Xu et al., 2024 | Health Expectancy | ✓ |  |  |  |  |  | ✓ |  |
| Yu et al., 2024 | WHO Intrinsic Capacity | ✓ | ✓ | ✓ |  |  |  |  |  |
| Zhou and Wang et al., 2024 | WHO Intrinsic Capacity | ✓ | ✓ | ✓ |  |  | ✓ |  |  |

**Table 2** Measurement of socioeconomic indicators and number of studies including the respective operationalisation.

| **SES indicators** | **N of studies** | **References** |
| --- | --- | --- |
| **Educational measured** | 43 |  |
| 1. Educational attainment |  |  |
| Three categories |  |  |
| - Primary, secondary, and post-secondary | 7 | Amaia Calderón-Larrañaga/ Boris Cheval/ Esther García-Esquinas/ Lisa Harber-Aschan/ Ziting Huang (respondents)/ Yu-Tzu Wu (a)/ Yu-Tzu Wu (b)/ |
| - Illiterate, high school, and above high school | 6 | Hui Chang/ Javier de la Fuente(b)/ Huiying Liu (respondents)/ Li-Fan Liu/ Olivia S. Malkowski/ Ruby Yu/ |
| - Lower secondary, upper secondary or postsecondary non-tertiary level of education, and first stage of tertiary education or higher | 2 | Haomiao Li/ Kim Qinzi Xu/ |
| More than 3 categories | 10 | Joan Domènech-Abella/ Almar A L Kok (a)/ Wentian Lu/ Sara J. McLaughlin/ Rachel A. Pruchno/ Aaron Salinas-Rodríguez/ Yafei Si (respondents)/ Christine Stephens/ Erwin Stolz/ Christine M. White/ |
| Binary variable | 12 | Julie E. Byles/ Francisco Félix Caballero/ Yuanyan Chen/ Javier de la Fuente (a)/ Yanfei Guo/ Ziting Huang (parents)/ Huiying Liu (parents)/ Dieta Nurrika/ Collin F. Payne/ Yafei Si (parents)/ Xu Wang/ Yaru Zhou/ |
| 2. Years of education |  |  |
| Continuous | 8 | Carmen Arroyo-Quiroz/ Ottavia Eleonora Ferraro/ Franque Grimard/ Hui-Chuan Hsu/ Almar A. L. Kok (b)/ Suzanne Meeks/ Xiao Xu/ |
| Categorical variable | 1 | Theodore D. Cosco/ |
| 3. Numbers of books in home | 2 | Boris Cheval/Christine Stephens/ |
| 4. Age left school | 1 | Elise Whitley/ |
| **Income/wealth** | 36 |  |
| 1. Personal income |  |  |
| - Specific values as cut-off points | 1 | Li-Fan Liu/ |
| ­- Tertile, quartile, or quintile groups | 1 | Elise Whitley/ |
| 2. Household income |  |  |
| - Specific values as cut-off points | 0 |  |
| ­- Tertile or quintile groups | 4 | Almar A L Kok (a)/ Wentian Lu/ Erwin Stolz/ Xiao Xu/ |
| - Continuous | 3 | Linda Juel Ahrenfeldt/ Joan Domènech-Abella/ Christine M. White/ |
| - No specific classification methodology reported | 1 | Collin F. Payne/ |
| 3. Household wealth |  |  |
| - Specific values as cut-off points | 0 |  |
| - Quartile or quintile groups | 11 | Francisco Félix Caballero/ Javier de la Fuente(b)/ Ziting Huang/ Wentian Lu/ Olivia S. Malkowski/ Aaron Salinas-Rodríguez/ Erwin Stolz/ Yu-Tzu Wu (a)/ Yu-Tzu Wu (b)/ Kim Qinzi Xu/ Xiao Xu/ |
| - Binary variable | 1 | Javier de la Fuente (a)/ |
| - Continuous | 1 | Linda Juel Ahrenfeldt/ |
| - No specific classification methodology reported | 1 | Collin F. Payne/ |
| 4. Perceived income/wealth measures |  |  |
| - Perceived income adequacy, confidence in future finances, satisfaction with economic/financial status, financial strain, difficulty in managing income. | 14 | Julie E. Byles/ Amaia Calderón-Larrañaga/ Yuanyan Chen/ Boris Cheval/Yanfei Guo/ Lisa Harber-Aschan/ Ziting Huang/ Huiying Liu/ Yafei Si/ Christine Stephens/ Christine M. White/ Elise Whitley/ Xu Wang/ Ruby Yu/ |
| 5. Other |  |  |
| - Pension | 2 | Hui Chang/ Yuanyan Chen/ |
| - Monthly per capita expenditure | 1 | Dieta Nurrika/ |
| - Annual household consumption expenditure | 2 | Huiying Liu/ Collin F. Payne/ |
| - Log per capita assets | 1 | Franque Grimard/ |
| - Economic hardship | 1 | Else Foverskov/ |
| - Household per capita expenditure | 1 | Haomiao Li/ |
| - Equivalized income decile | 1 | Mengling Cheng/ |
| **Occupational position** | 22 |  |
| 1. Occupational level |  |  |
| - Three categories | 9 | Hans Bosma/ Boris Cheval/ Joan Domènech-Abella/ Lisa Harber-Aschan/ Huiying Liu (respondents)/ Olivia S. Malkowski/ Erwin Stolz/Yu-Tzu Wu (b)/ Kim Qinzi Xu/ |
| - Five categories | 1 | Christine Stephens/ |
| - Five categories including “never had job” | 1 | Almar A L Kok (a)/ |
| - Six categories | 2 | Christine M. White/ Elise Whitley/ |
| 2. Manual vs. non manual | 1 | Amaia Calderón-Larrañaga/ |
| 3. Agricultural vs. nonagricultural | 3 | Yuanyan Chen/ Yanfei Guo/ Collin F. Payne/ |
| 3. Employment status (employed vs. unemployed) | 3 | Francisco Félix Caballero/ Rachel A. Pruchno/ Xu Wang/ |
| 4. Other | 3 | Ziting Huang/ Huiying Liu (parents)/Almar A. L. Kok (b)/ |
| **Childhood variables** |  |  |
| 1. parental education | 10 | Hans Bosma/ Ziting Huang/Yanfei Guo/ Huiying Liu/ Almar A. L. Kok (b)/ Collin F. Payne/ Rachel A. Pruchno/ Yafei Si/ Christine Stephens/ Yu-Tzu Wu (b)/ 11 |
| 2. parent occupation | 13 | Hans Bosma/ Yuanyan Chen/ Boris Cheval/ Else Foverskov/ Yanfei Guo/Lisa Harber-Aschan/ Ziting Huang/ Huiying Liu/Almar A. L. Kok (b)/ Collin F. Payne/ Christine Stephens/Elise Whitley/ Yu-Tzu Wu (b)/ |
| 3. income/wealth in childhood | 4 | Hans Bosma/ Ziting Huang/ Yafei Si/ Xu Wang/ |
| 4. other | 1 | Franque Grimard/ |
| **Other variables** |  |  |
| **1.Housing variables (Housing tenure, housing quality,** **overcrowding)** | 5 | Boris Cheval/ Lisa Harber-Aschan/ Christine Stephens/Elise Whitley/ Ruby Yu |
| **2. Hukou** | 2 | Collin F. Payne/ Yafei Si |
| **3. Urban** | 7 | Hui Chang/ Yuanyan Chen/ Joan Domènech-Abella/ Haomiao Li/ Collin F. Payne/ Yafei Si/ Xu Wang/ |
| **4. Neighbourhood environment** | 2 | Huiying Liu / Yafei Si/ |

**Table 3** Heterogeneity of SEP operationalisation and methodologies included in studies

| **Study** | **Education** | **Income/Wealth** | **Occupation** | **Others** | **Model and adjustments** |
| --- | --- | --- | --- | --- | --- |
| Ahrenfeldt et al., 2021 |  | Continuous: Household income and Household wealth |  |  | Structural equation modelling   - Model 2 was adjusted for age group, European region, wave, and marital status. - Model 3 was further adjusted for employment |
| Arroyo-Quiroz et al., 2020 | Continuous: Years of education |  |  |  | Logistic regression; Cox proportional hazards models   - adjusted for age, sex, education, marital status, smoking, alcohol consumption, physical activity, self-perceived depression, BMI category, follow-up time, and parental longevity. |
| Bosma et al., 2007 |  |  | - High - Intermediate - Low |  | Linear regression analyses   - adjusted for age, sex, baseline level of functioning, early socioeconomic and developmental factors, and intellectual abilities. |
| Byles et al., 2019 | - <Higher school certificate - ≥Higher school certificate | Difficulty in managing income:   - Easy/not too bad - Difficult some/all time |  |  | Repeated measures latent class analysis; Multivariable multinomial logistic regression   - Adjusted for marital status, income management, education, smoking, BMI, exercise, and social support. |
| Caballero et al., 2017 | - No qualification - Some formal education | Household Wealth: Quintiles groups | - Not in work - Being in work |  | Mixed-effects multilevel regression   - adjusted for gender, age group, household wealth, formal education, marital status, falls, smoking status, alcohol consumption, physical activity, employment, and size of social network. |
| Calderón-Larrañaga et al., 2021 | - Elementary - High school - University or higher | Financial strain: yes/no | - Manual - Non-manual |  | Growth Mixture Models; Logistic regression   - adjusted for baseline age, sex, and all socioeconomic, psychosocial, and behavioural factors simultaneously. |
| Chang et al., 2023 | - Illiterate - Primary and below - Junior high school and above | Retirement pension: yes/no |  | Residence:   - Rural - Urban | Growth mixture modelling; Multinomial logistic regression analysis   - Adjusted for age (categorical), gender, marital status, education level, residence, smoking, drinking, self-rated health, life satisfaction, retirement pension. |
| Chen et al., 2024 | - Any - None | Pension: yes/no  financial sufficiency: adequate/not | father's occupation:   - Non-agricultural - Agricultural   primary occupation before 60:   - Non-agricultural - Agricultural | birth type:   - Urban - Rural   current residence:   - Urban - Rural | Continuous-time multi-state models; multinomial regression   - Adjusted for age, gender |
| Cheng et al., 2023 |  | Continues: Equivalized income decile |  |  | Poisson growth curve modelling   - Adjusted for wealth decile, education level, gender, region of residence (urban/rural), current marital status, current working status, and household size. |
| Cheval et al., 2019 | number of books at home:   - 0-10 - More books at home   educational attainment:   - Primary - Secondary - Tertiary | satisfaction with household income:   - With great difficulty - With some difficulty - Fairly easily - Easily | occupational position of main breadwinner   - First and second - Higher skills levels   main occupational position   - Low skill - High skill - Never worked | overcrowding   - More than one person per room - Less than one person per room   housing quality   - Absence of either fixed bath, cold running water supply, hot running water supply, inside toilet or central heating - Presence of above | Linear growth curve models   - adjusted for age at baseline, birth cohort, adverse childhood experiences, childhood health problems, participant attrition, and country of residence. |
| Cosco et al., 2017 | - 0-9 years of full-time education - 10-11 years of full-time education - ≥12 years of full-time education |  |  |  | Growth Mixture Modelling; Ordinal logistic regression   - Adjusted for age, sex, marital status, and occupational status. |
| Domènech-Abella et al., 2017 | - Incomplete primary school - Primary school - Lower secondary school - Upper secondary school - College/university | Continuous: total household income | - Skill Level 1 - Skill Level 2 - Skill Level 3 | Urbanicity:   - Rural - Urban | Linear regression models   - adjusted for all sociodemographic variables simultaneously (sex, age, education, occupation, marital status, urbanicity). |
| Ferraro et al., 2021 | Continuous: Years of education |  |  |  | Group-Based Trajectory Model; multinomial logistic model   - Not applicable |
| Foverskov et al., 2020 |  | Accumulated Economic hardship (EH):  0, 1, 2, 3, 4 or more years in EH  EH trajectories:   - Low probability - Declining probability - Rising probability - High probability |  |  | Linear regression models   - adjusted for sex, age group, original cohort, long-term parental unemployment/financial problems, educational level, and baseline income |
| de la Fuente et al., 2018 | having a degree or certificate: yes/no | Household wealth:   - 1st-2nd quintile - Other quintiles |  |  | Growth Mixture Model; Multinomial logistic regression model   - adjusted for age, health at baseline, gender, household wealth, number of chronic conditions, and (for HRS) ethnicity. |
| de la Fuente et al., 2019 | - No education - Medium education - High education | Household wealth: Quintiles groups |  |  | Bayesian mixed-effects multilevel models   - The models included linear and quadratic effects of age and birth year, education level, household wealth quintile, and all interactions between these variables. |
| García-Esquinas et al., 2019 | - Primary or less - Secondary - University |  |  |  | Linear mixed models   - adjusted for sex, educational level, baseline diet score, and changes over time in smoking, alcohol consumption, physical activity, sedentary behaviour, and body mass index. |
| Grimard et al., 2010 | Continuous: Years of education | Continuous: Log per capita assets |  |  | Probit model   - adjusted for age, marital status, location size dummies, and survey year. |
| Guo et al., 2025 | Parental education:   - No formal education - Formal education | Family financial situation:  Yes/no | Parental occupation:   - Agriculture - Non- agriculture | Childhood SEP:   - Low - Moderate - High   Adult SEP: (15 items included in PCA)   - Low - Moderate - High | Group-based trajectory models; Causal mediation analysis; Multinomial logistic regression model   - Adjusted for age, gender, place of residence (rural/urban), tobacco use, alcohol consumption, physical activity, region of China, household wealth per capita. |
| Harber-Aschan et al., 2020 | - Elementary - Secondary - Post-secondary or university | childhood financial strain/lack of financial assets/financial strain:  yes/no | Parental occupation/ Participants’ occupation:   - Manual - Non-manual - Professional | Homeownership:   - Owning property - Rentals or other forms of housing | Linear Mixed Models   - Adjusted for age, sex, civil status, migrant status, smoking, alcohol use, BMI, and depressive symptoms. |
| [Hsu](javascript:;) et al., 2012 | Continuous: Years of education |  |  |  | Multiple group-based trajectory analysis; Multinomial logistic regression   - Adjusted for age, gender, education, marital status, and residence. |
| Huang et al., 2025 | - Primary - Secondary - Tertiary   Parents’ education:  0/1 | Financial difficulties during childhood: 0/1  Household wealth:  Tertiles group | Parents’ occupation:  0/1 |  | Linear mixed models   - Adjusted for age, sex, marital status, and follow-up years. |
| Kok et al., 2016 | - Elementary not completed - Elementary - Low - Intermediate - High | Household income:  Quintiles groups | - Elementary - Low - Medium - High - Never had a paid job |  | Latent Class Growth Analysis; Multivariate regression models   - adjusted for baseline age and sex. |
| Kok et al., 2017 | Continuous: 5-18 years of education |  | father's occupational prestige and  occupational prestige:  coded into a prestige scale (range 13-82) |  | Path analysis   - adjusted for baseline age. Effects that did not differ by gender were also adjusted for gender. |
| Li et al., 2022 | - Less than lower secondary - Upper secondary - Vocational | Household per capita consumption:   - Low - Low-to-middle - Middle - High |  | Residence:   - Rural - Urban | Latent Growth Mixture Model; Multinomial Logistics Regression Model   - adjusted for socioeconomic background (age, gender, marital status, education, consumption), family characteristics (e.g., care to grandchildren, contact with children), and lifestyle factors (e.g., alcohol, smoking, social participation). |
| H Liu et al., 2023 | Education attainment:   - Illiteracy - Elementary school - Higher than elementary school   Parents’ education:   - Both illiterate - At least one parent literate | family economic status in childhood:   - Poor - A little better than others - Good   annual household expenditure:   - Low - Medium - High   self-rated economic status: yes/no | Parents’ occupation   - Both working in agriculture - At least one parent not working in agriculture   primary occupation in adulthood:   - Farming/unpaid domestic work - Worker/clerk/service worker - Professionals/managers | Life-course SES disadvantages (eight groups)  Community environment resources (continuous) | Multilevel growth modelling; Marginal structural models   - Adjusted for demographic characteristics (gender, marital status, birth cohort), early-life health (serious diseases in early life), health behaviours (smoking, alcohol consumption, physical exercise), urban-rural residence, presence of elder care centre, provision of old-age allowance. |
| Li Liu et al., 2016 | - Illiterate - Elementary school - High school | Economic status:   - <60000 NTD - 60000-120000 - 120000-240000 - >240000 |  |  | Latent Class Analysis; Generalized Estimating Equation   - adjusted for age, sex, wave (time), marital status, religion, location, education, income, smoking, alcohol consumption, and social participation. |
| Lu et al., 2021 | - First stage of tertiary or more - Post-secondary non-tertiary - Upper secondary education - Lower secondary education - Primary education or less | Household wealth:  Quintiles groups  Household income:  Quintiles groups |  |  | Multi-level modelling   - adjusted for other SEP indicators, age, age-squared, cohort, cohort-squared, gender, ethnicity, childhood self-rated health, father's occupation, own occupation, marital status, smoking, drinking, and relevant interaction terms. |
| Malkowski et al., 2023 | - No formal qualifications - Secondary or lower - At least some higher education | Household wealth: Quintiles groups | - Routine and manual occupations - Intermediate occupations - Higher occupations |  | Growth Mixture Modelling; Multinomial logistic regression   - adjusted for age, biological sex, and ethnicity. |
| McLaughlin et al., 2020 | - less than a high school (HS) diploma - HS diploma - Some college - A college or higher degree |  |  |  | Generalized Estimating Equations   - adjusted for demographic covariates (age, gender, marital status, wealth, race/ethnicity). |
| Meeks et al., 2001 | Continuous: years of education |  |  |  | Structural Equation Modelling   - The final reported models were unadjusted, initial models included age and gender, but these variables did not add significantly to the models and were removed to improve parsimony. |
| Nurrika et al., 2020 | - Below lower-secondary education - Lower-secondary education and above | Monthly per-capita expenditure:   - ≤ 43 USD - > 43 USD |  |  | Multivariate-adjusted logistic regression model   - adjusted for age, sex, region, area, and either monthly PCE (when education was the exposure) or education level (when PCE was the exposure). |
| Payne et al., 2022 | - Primary or less - More than primary | annual household income, annual household consumption expenditure, household wealth: continuous | - Non-agricultural - Agricultural |  | Cumulative logistic regression model   - adjusted for age, age squared, sex, and SES. |
| Pruchno et al., 2015 | 9-point scale, 1 = not high school graduate, 9 = professional degree |  | Working status: yes/no |  | Multinomial Logistic Regression   - adjusted for: Early life influences (age, gender, race, education, never-married status, incarceration, childlessness) and Midlife characteristics (being married, working, volunteering, smoking cigarettes, alcohol consumption, Body Mass Index (BMI), cardiovascular exercise, social support, religiosity). |
| Salinas-Rodríguez et al., 2024 | Educational level: Quintiles groups | Household wealth: Quintiles groups |  |  | Growth mixture models; Generalized linear models; Multinomial logistic regression   - Adjusted for age, marital status, having a paid job, health insurance, multimorbidity, physical activity, sedentary behaviour, tobacco use, alcohol consumption, and intake of fruits/vegetables. |
| Si et al., 2023 | Educational achievement:   - Illiterate - Primary - Middle school - High school - College and above   Parental education:   - Literate - Illiterate | Current family economic status:  Quintiles groups  family economic status during childhood:   - Low - Average - High |  | Neighbourhood environment:  Sum score of 4 items (range 0-4)  Urban residence:   - Rural - Urban | Multivariable linear regression   - Adjusted for current socioeconomic factors (family economic status, education, urban residence, urban hukou), demographic factors (age, gender, marital status), and lifestyle factors (tobacco consumption, alcohol consumption, presence of chronic diseases). |
| Stephens et al., 2022 | - No formal qualification - Secondary qualification - Post-secondary qualification - University degree   number of books available in the household:   - 0–10 books - 11–25 books - 26–100 books - 101–200 book - more than 200 books | financial history: five items  income met  needs for necessities:   - Not enough - Just enough - Enough - More than enough   Financial hardship:  total number of periods  when income was ‘not enough’ for everyday needs. | main breadwinner's occupational grade (childhood) & occupational grade:   - No main breadwinner - Elementary - Skilled - Associate - Manager | Housing quality:   - > 1.5 people per room - < 1.5 people per room | Latent growth curve analysis; Mediation analysis   - Adjusted for age |
| Stolz et al., 2017 | - Primary - Lower secondary - Upper secondary - Postsecondary | Household income: Quartiles groups  Household wealth: Quartiles groups | - Salariat - Intermediate - Working class |  | Growth curve models; hierarchical linear modelling   - adjusted for age, sex, living alone, birth cohort, non-response (attrition), and death of respondent. |
| Wang et al., 2024 | - Elementary school or below - Middle school or above | Family financial status in childhood:   - Worse off than others - Same as others - Better off than others | - Employed - Not employed | Residence:   - Urban - Rural | Latent Class Growth Analysis; Multinomial logistic regression   - Adjusted for: Age, Gender, Education level, Work status, Current residence, Depression, Number of chronic diseases, social activity, Friendship in childhood, Adverse childhood events, and Family financial status before age 17. |
| White et al., 2015 | Eleven possible levels, including "No formal schooling", "Some primary school", up to "Master's degree or PhD" | Perceived income adequacy:   - Totally inadequate - Not very well/some difficulty - Adequately - Very well   Life satisfaction with finances:   - Not happy - Happy - Very happy   Household income: continuous | - Unskilled - Semiskilled - Farmers - Skilled - Technicians and middle management - Professionals |  | Logistic regression   - adjusted for age and gender. |
| Whitley et al., 2018 | Age left school:   - At or before leaving age - Beyond leaving age Full school education | Income:  Quintiles groups  Ease of making ends meet:   - Difficult - Moderate - Easy   Confidence in future finances   - Very insecure - Fairly insecure - Fairly secure - Very secure | Parental occupation:   - I - II - III non-manual - III manual - IV - V | Housing tenure and car ownership:   - No tenure/no car - No tenure/car - Tenure/no car - Tenure/car | least squares regression; logistic regression   - adjusted for gender and age-cohort. |
| Wu et al., 2020 | - Primary education or less - Secondary education - Tertiary education | Household wealth:  Quintiles groups |  |  | multilevel modelling   - adjusted for age, sex, and cohort study. |
| Wu et al., 2025 | - Primary - Secondary - Tertiary | Household wealth:  Quintiles groups | - Low - Middle - High | Childhood SEP：   - Low - Middle - High | Causal mediation analysis; Linear regression modelling   - adjusted for age and sex. |
| X Xu et al., 2014 | Continuous: years of schooling | Household income &  Household net worth:  Quartiles groups (two higher quartiles combined as the reference group) |  |  | multinomial logistic regression   - adjusted for gender, race/ethnicity, baseline age, baseline self-rated health, baseline number of chronic diseases, mortality, and attrition. |
| K Xu et al., 2024 | - No year 12/no post-secondary - Year 12 or post-secondary - Tertiary | Household wealth:  Tertiles groups | - Low - Middle - High |  | multinomial logistic regression.   - Adjusted for transition probabilities included age, age², sex, and SEP measures, as well as interactions between age, sex, and SEP. |
| Yu et al., 2024 | - Mo education - Primary - Secondary or above | Financial assistance status:   - Recipient - Non-recipient   Perceived financial adequacy:  A 5-point scale from very inadequate to very adequate |  | Housing type:   - Public - Subsidized - Private | Cox proportional hazards models   - Adjusted for age, sex, marital status, educational attainment, financial assistance status (CSSA), housing type, perceived financial adequacy, number of chronic diseases (0, 1, 2+), and self-rated health. |
| Zhou and Wang et al., 2024 | - Primary school and below - Junior high school and above |  |  |  | Logistic regression analysis   - Adjusted for age, education level (primary school and below), marital status (widowed, others vs. married), and baseline IC score |
